# Whole Genome Sequencing in Adolescent Idiopathic Scoliosis Cohort Indicates Polygenic Disease Involving Multiple Biological Pathways

**DOI:** 10.1101/2025.01.23.25321049

**Authors:** Islam Oguz Tuncay, Eun Kyoung Lee, Anxhela Gustafson, Yoonsuh Lee, Dawoon Jung, June-Young Koh, Wonchul Lee, Sangmoon Lee, Kamran Shazand

## Abstract

Adolescent idiopathic scoliosis (AIS) is the most common nondegenerative spinal abnormality. Research indicates a strong correlation between genetics and AIS, with heritability estimates of 87.5%. However, the rarity of shared causative genes among patients, and the difficulty of replication between studies suggest that AIS is a highly complex polygenic disease. In this study, we utilized whole-genome sequencing (WGS) to comprehensively explore the genetic landscape of 119 AIS patients from 103 families. We implemented an automated WGS analysis pipeline powered by RareVision^TM^ consisting of automated algorithms and manual curation. We identified clinically relevant candidate variants in 20/119 (16.8%) patients and potentially relevant strong or moderate candidate variants in another 73/119 (61.3%) patients. Candidate variants included coding and noncoding point mutations, along with structural variants and large indels, showing the utility of WGS. Candidate genes included AIS-associated genes (e.g. *CHD7*, *COL11A1/2*, *FBN1/2*, *HSPG2*, *KIF7*), as well as genes associated with other musculoskeletal and developmental syndromes where scoliosis is a known symptom (e.g. *RYR1*, *GJB2*, *MYH2*, *MYH7*). Association analysis showed 4 known AIS single nucleotide polymorphisms (rs12946942, rs10756785, rs3904778, rs7633294) also correlated with AIS in our cohort. Finally, through gene set enrichment analysis, we were able to identify 3 gene clusters involved in skeletal muscle contraction, extracellular matrix composition, and gene expression regulation. In summary, through scalable WGS-based familial testing we were able to (1) find clinically relevant genetic variations in the majority of our patients and (2) create a large dataset that allowed us to identify biological pathways relevant to AIS etiopathogenesis.

## INTRODUCTION

Adolescent idiopathic scoliosis (AIS) is the most common nondegenerative spinal abnormality with a prevalence of 1−4%.^1–3^ It is diagnosed when a three-dimensional spinal deformity measured by the Cobb angle exceeding 10° of magnitude in the coronal plane.^4^ Severe progressive curvatures can lead to serious complications such as cardiac or respiratory compromise, back pain, and other degenerative disease, as well as psychological issues related to cosmetic deformity, which can have a long-term impact on the patient’s quality of life.^5^ For mild scoliosis, brace therapy is often combined with physiotherapy, and for severe individuals, surgical treatments are required.

Despite growing research for decades, the etiopathogenesis of AIS remains unclear. Several hypotheses of AIS etiology involving genetics, central nervous system, biomechanics, metabolic pathways, spinal cord growth, endocrine factors, sex hormones, bone metabolism, and epigenetics have been proposed.^6,7^ Epidemiological studies have identified high heritability and marked sexual dimorphism in AIS, with significant female predominance.^1^ Multiple literatures support the genetic foundation of AIS, with sibling recurrence risks reported to be 17.7%, heritability estimates of approximately 87.5%,^8^ and odds ratio (OR) for developing AIS being 1.5-fold higher for the participants whose mother had scoliosis.^9^ Nevertheless, the rarity of causative genes that have been clearly identified as ‘AIS genes’ and the lack of replication of certain variants between genetic analysis in different study cohorts suggest that AIS is a highly complex polygenic disease that results from the interaction of multiple gene loci and the environment.^10–12^

With the advancement of next-generation sequencing, molecular genetics diagnostic methods using targeted sequencing or whole exome sequencing (WES) have been widely utilized. Most AIS genetic researches have mostly utilized targeted sequencing, WES, and single nucleotide polymorphism (SNP)-based genome-wide association studies (GWAS).^13–28^ However, mutations in regulatory or intronic regions, difficult-to-sequence repetitive sequences, and the presence of novel genes are challenging to uncover with targeted sequencing or WES, and the complex genomic changes including structural variations (SVs) are difficult to identify with SNP-based GWAS. Whole genome sequencing (WGS), covering the entire genome allows for a more comprehensive exploration of the extended genetic landscape. The clinical utilization of WGS has been limited due to high cost and excessive data, however, a fall in the costs of high-throughput sequencing and advances in bioinformatic analyses have increased the potential for WGS to be used in clinical practice.^29^

In this study, we comprehensively explored the genetic landscape in 103 probands with AIS utilizing WGS. For WGS analysis, we implemented an automated WGS analysis pipeline powered by RareVision^TM^ consisting of automated algorithms and manual curation by specialists of genomic medicine. The main goal of this study was to find new candidate pathways and functional categories involved in scoliosis by analyzing a large cohort of AIS patients.

## MATERIAL AND METHODS

### Study Subjects

This is a retrospective analysis of patients with AIS at the Research for Precision Medicine study at Shriners Hospitals for Children – The Genome Institute. The study was approved by the Institutional Review Board of Shriners Hospitals for Children (IRB approval number: 9-21-2022) and adhered to the tenets of the Declaration of Helsinki. All patients underwent an orthopedic examination to confirm the idiopathic nature of the scoliosis. Patients with congenital scoliosis and possible conditions that cause neuromuscular scoliosis including cerebral palsy, Duchenne muscular dystrophy, myelomeningocele, spinal muscular atrophy, or Friedrich ataxia were excluded from the study. The radiographic diagnostic of scoliosis relied on the presence of a lateral curve to the spine greater than 10° with vertebral rotation.^30^ The demographic data and clinical phenotypes were retrieved from the electronic medical records. A pedigree investigation was conducted for all patients, and the principle of performing genetic testing for all available parents was followed. Genome sequencing was performed as singleton, duo, and trio sequencing depending on availability of biological parental samples. All individuals provided informed consent prior to the genetic analyses.

### Whole-Genome Sequencing and Bioinformatics Analysis

To obtain genomic DNA, peripheral blood samples were collected from probands with or without their parents. Shriners Genomics Institute performed the entire genome sequencing process, and analysis and interpretation were performed using the RareVision™ system (Inocras Inc., San Diego, CA, USA). Genomic DNA was extracted from blood samples using the Allprep DNA/RNA kits (Qiagen, Venlo, Netherlands). DNA libraries were prepared using TruSeq DNA PCR-Free Library Prep Kits (Illumina) and sequenced on the Illumina NovaSeq6000 platform (Illumina) with an average depth of coverage of 30×. The obtained genome sequences were aligned to the human reference genome (GRCh38) using the BWA-MEM algorithm. PCR duplicates were removed using SAMBLASTER. The initial mutation calling for base substitutions and short indels was performed using HaplotypeCaller2 and Strelka2, respectively. Structural variations were identified using Manta. Variants were filtered and annotated. Additionally, to focus on rare variants, we selected variants with minor allele frequency (MAF) <0.01 in 1000 Genomes Project (1000GP),^31^ The Genome Aggregation Database (gnomAD),^32^ and The Exome Aggregation Consortium (ExAC).^33^ Pathogenicity was defined based on the clinical information and genetic variant information of the test subjects, following the guidelines published by the American College of Medical Genetics and Genomics (ACMG),^34^ ClinVar annotations and functional prediction scores: variants reported to be either (1) pathogenic, (2) likely pathogenic, or (3) uncertain significance. The pathogenicity prediction was further enhanced by using in-house-developed software that automatically integrates updated databases for more accurate analysis.

### Candidate Variant Prioritization

To prioritize candidate variants, we considered (1) previously reported associations between the gene and AIS, (2) predicted effect of the mutation, and (3) familial segregation. For (1), we employed a filter using Human Phenotype Ontology (HPO) term “Scoliosis”, and conducted a gene-based review in OMIM and PubMed to sort the candidate genes into the following categories: Known AIS genes (K-AIS) includes 12 genes with strong evidence for AIS association in previous studies (*ADGRG6*/*GPR126*,^35^ *AKAP2*,^36^ *BNC2*,^37^ *CHD7*,^38,39^ *COL11A1*,^40^ *COL11A2*,^41^ *FAT3*,^42^ *FBN1*,^43^ *FBN2*,^43^ *HSPG2*,^44^ *KIF7*,^45^ *LBX1*,^46^ and *POC5*^47^). Autosomal dominant/recessive syndromic AIS genes (S-AD-AIS/S-AR-AIS) includes genes known to be associated with an OMIM-listed genetic syndrome where scoliosis is a reported symptom or common comorbidity. Genes with possible association to AIS (Assc-AIS) includes genes with previous evidence for association with AIS, other skeletal abnormalities, or other AIS-associated genes. All remaining genes were considered “NKA” – no known association.

For (2) predicted effect of the mutation, we grouped structural, frameshift, start loss, stop gain (nonsense), stop loss, and splice-site variants as protein truncating variants (PTVs). Missense variants that were reported as pathogenic/likely pathogenic in ClinVar, and/or predicted to be pathogenic/likely pathogenic by the ACMG guidelines were grouped as P/LP. Variants that were predicted to be benign/likely benign based on the ACMG guidelines, variants that were predicted to be benign/tolerated/likely benign by the majority of available predictive tools and databases, and variants that were marked as benign/likely benign in ClinVar were grouped as B/LB. The remaining candidate variants were considered variants of uncertain significance, or “VUS”.

For (3) familial segregation, variants categorized based on family type and mode of inheritance. Our cohort included trios, duos, and singletons. Note that the phenotype information was unavailable for some participating parents. These cases were evaluated based on the number of parents with known phenotype. Following modes of inheritance were considered: *De novo* variants were absent in both unaffected parents, and heterozygous in the proband. Inherited homozygous variants were heterozygous in both unaffected parents, and homozygous in the proband. Inherited heterozygous variants were absent in the unaffected parent, and heterozygous in the affected parent and the proband. Compound heterozygous variants were defined as a two different inherited heterozygous or *de novo* variants present in the same gene in *trans*. For unknown parental genotype and phenotypes, all possible options were considered, and the candidate variants were evaluated for each combination to select the strongest option. Variants that did not fit the criteria for *de novo*, heterozygous, homozygous, or compound heterozygous based on available parental genotype and phenotypes were considered non-segregating. Based on these criteria, the candidate genes were sorted into categories as described in the Table 1 as follows: 1) solved case, 2) nearly solved case, requires missing parent genotype, 3) case with strong candidate variant, 4) case with moderate candidate variant.

**Table 1.**
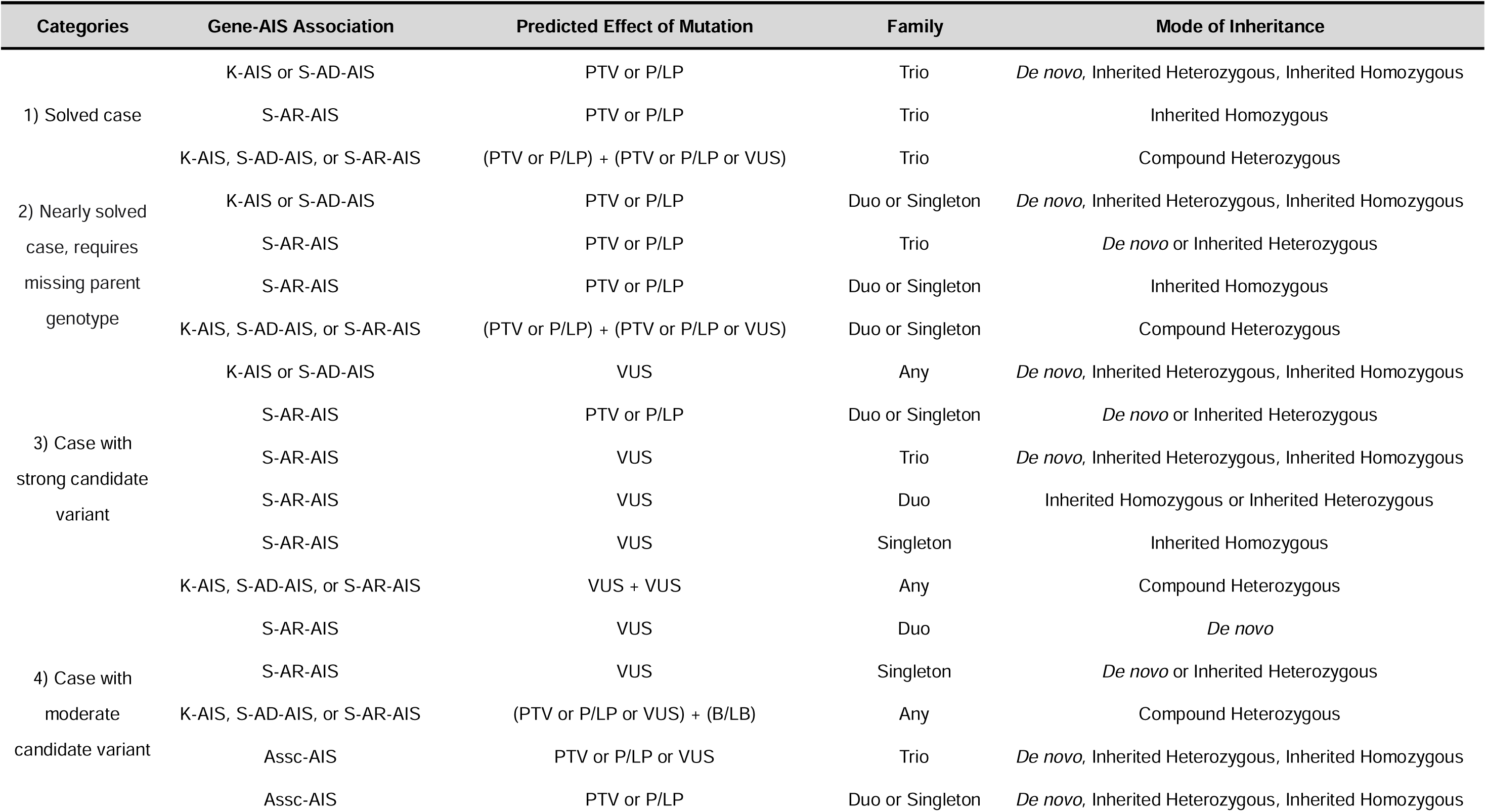
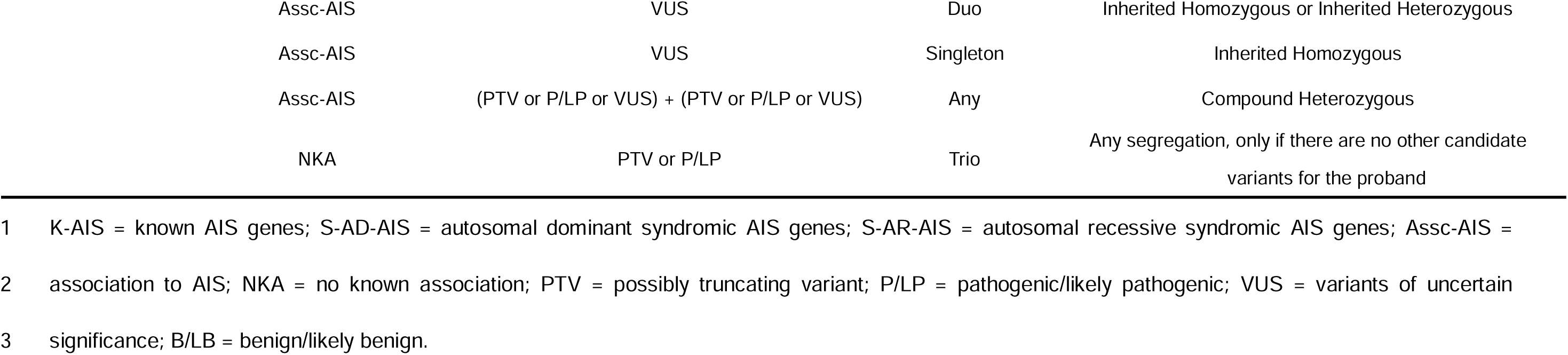
Candidate variant prioritization and definition of categories of cases with adolescent idiopathic scoliosis (AIS)

### Principal Component Analysis

Principal component analysis (PCA) was carried out in PLINK version 1.90b6.11^48,49^ using Phase 3 1000GP data.^31^ VCF files were converted into PLINK format. Autosomal variants were filtered for Hardy-Weinberg equilibrium (*p* < 0.001), MAF >5%, and maximum missing genotype rate of 25%. Proband and 1000GP cohort data were then merged and pruned to remove variants with MAF <10%, missing genotype rate greater than 5%, and pruned for linkage disequilibrium (LD) using PLINK –indep-pairwise 50 5 0.15. Triallelic and palindromic variants were also removed. PCA was run in PLINK using the –pca flag and the first two principal components were plotted in R.

### Gene Ontology Analysis

Genes that include pathogenic rare variants and AIS-related genes identified in the sequencing data were selected as candidate genes. Gene set overrepresentation analyses were performed on MAF < 0.05 gene lists with duplicate genes removed from the combined list from each family. We performed a molecular pathway analysis using the Kyoto Encyclopedia of Genes and Genomes (KEGG)^50^ to determine how candidate genes are networked and enriched within the pathways. Gene Ontology (GO)^51,52^ terms derived from cellular component and molecular function were also annotated to figure out protein-protein interactions. GO enrichment accessible at https://geneontology.org/ using GO enrichment analysis by PANTHER. The significance of an enrichment result was set at false discovery rate (FDR) <0.05 for all functional annotations. The same input gene lists as used for GO enrichment analysis were used in Enrichr, a gene enrichment software developed by the Ma’ayan laboratory^53^ accessible at https://maayanlab.cloud/enrichr-kg. We report results from the 2021 GO term Biological Process and 2021 KEGG Human pathways.

### SNP Association Analysis

The population of each proband was inferred by k-means clustering (KNN). Briefly, KNN was run on the first 3 principal components from the PCA output, using 1000GP data as the training set, and assuming k=5 for the 5 superpopulations in 1000GP (African or AFR, Admixed American or AMR, East Asian or EAS, European or EUR and Southeast Asian or SAS). For both probands and 1000GP samples, only the individuals with AMR or EUR backgrounds were included in the association analysis due to the rarity of other population groups in our cohort. An allelic test of association was performed for 15 SNPs previously associated with AIS^7^ using PLINK version 1.90b6.

## RESULTS

### Baseline Characteristics

The cohort comprised 204 samples from 103 families, consisting of 103 AIS probands and 101 relatives (16 singletons, 65 mother-only duos, 8 father-only duos, and 14 trios). Among the 103 probands, 81 patients (78.6%) were female, whereas 22 (21.4%) were male and 87 patients (84.5%) were idiopathic, whereas 16 (15.5%) were familial AIS with affected mother. Therefore, the analysis of candidate variants included 119 AIS patients (103 probands and 16 affected mothers) from 103 families. PCA with the 1000GP data shows that most of the participants including AIS probands and their parents have European and/or Admixed American ancestry (Figure 1).

**Figure 1.**
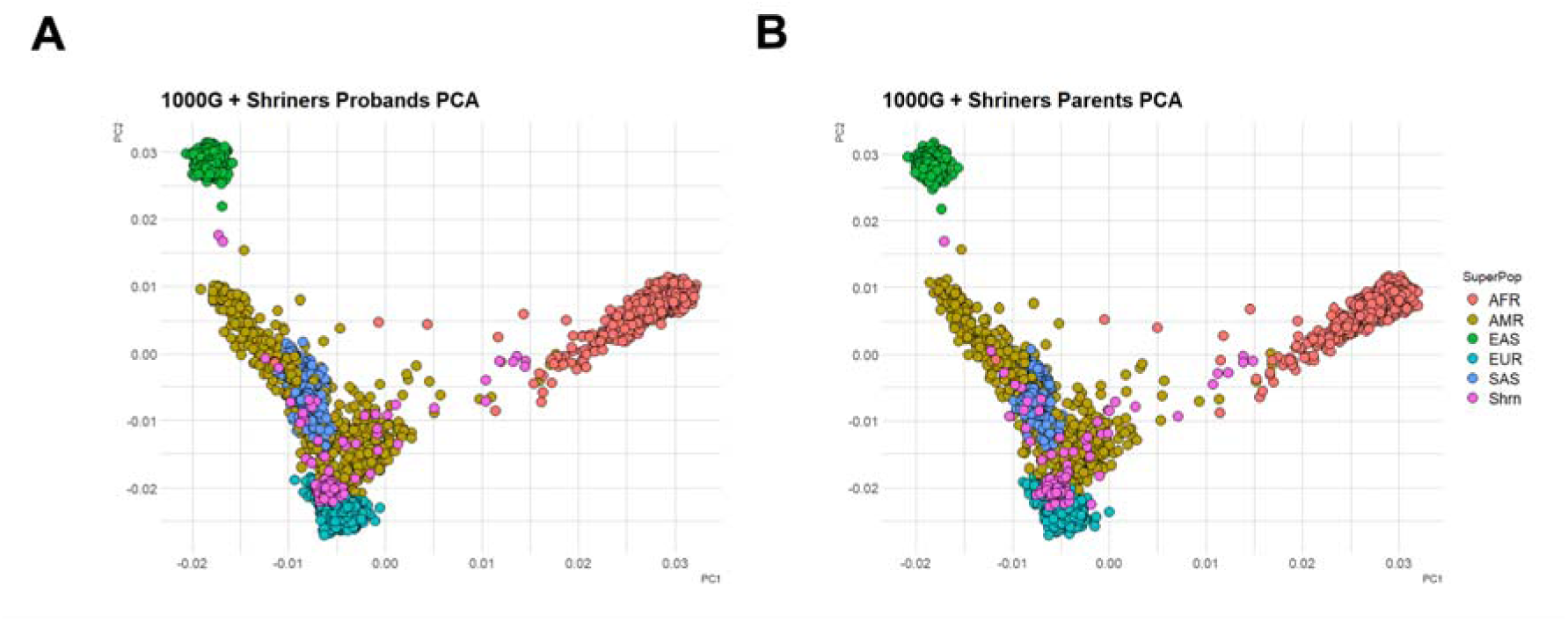
Principal component analysis of the adolescent idiopathic scoliosis (AIS) cohort. Principal component analysis (PCA) of the AIS cohort (**A**: probands, **B**: parents) combined with 1000GP. AIS cohort from Shriners Hospital are represented in pink. AFR, African; AMR, Admixed American; EAS, East Asian; EUR, European; SAS, South Asian; Shrn, Shriners.

### Candidate Variants

The average for mean genomic coverage was 40.7x and the average duplication rate was 7.9%. We identified clinically relevant candidate variants in 20/119 (16.8%) patients from 18/103 (17.5%) families, and potentially relevant strong or moderate candidate variants in another 73/119 (61.3%) patients from 61/103 (59.2%) families. WGS identified 137 genetic variants likely or highly likely to account for disease presentation. Figure 2A provides a summary of variant information identified across all probands. Missense variants constituted the majority of filtered variants (60.1%), followed by frameshift (16.7%), stop gain (8.0%), structural variations (7.2%), intronic variants (4.3%), and splicing variants (3.6%). A total of 27 variants were identified as ‘pathogenic’ or ‘likely pathogenic’ per ACMG guidelines. These genes include *ABCC9, ABL2, ACSF3, ALMS1, ALPK3, ANO5, ARID1B, ATP1A3, ATP7B, CUL7, CYP7B1, FBN1, GALT, GCDH, GNAI1, MFN2, MGAT5, NRCAM, NSD2, OTUD6B, PIGV, RECQL3, RNU4ATAC, SERAC1, SMARCA4,* and *TNPO3*.

**Figure 2.**
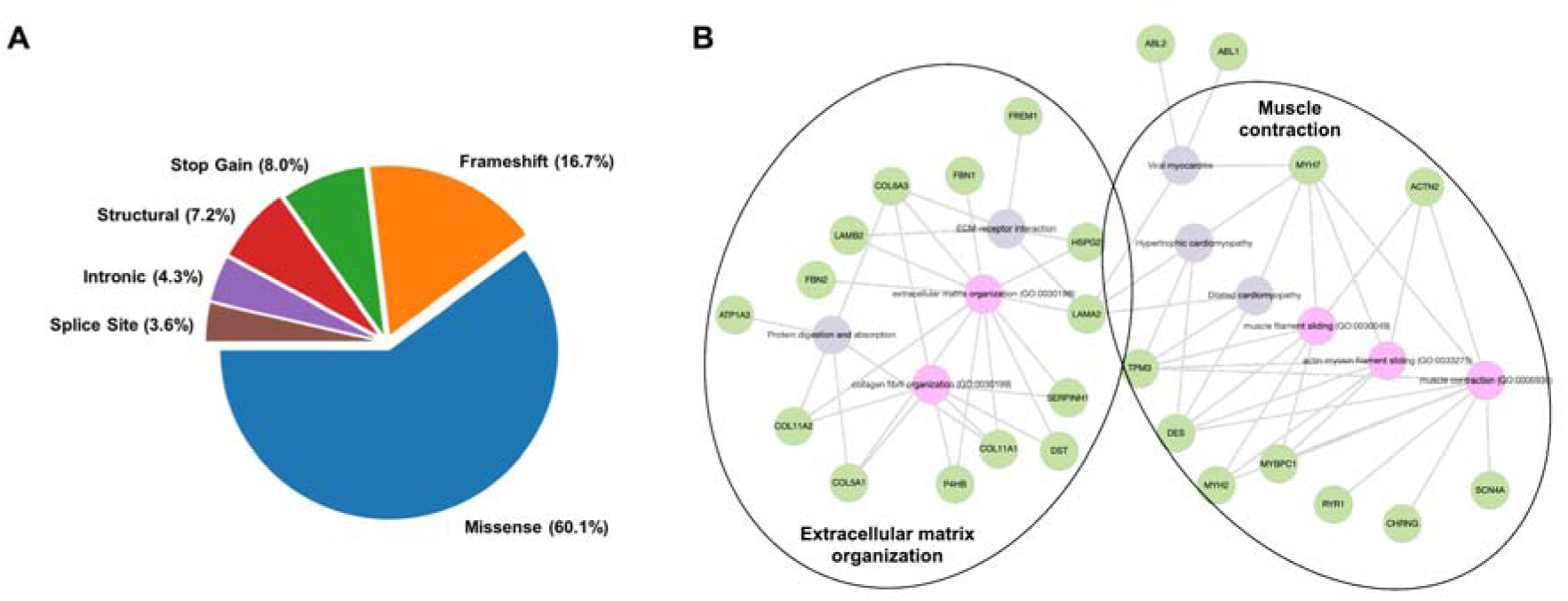
Summary information for total candidate gene found of the adolescent idiopathic scoliosis (AIS) cohort. **(A)** Pie-chart representing the repartition of category types of the variants found in the AIS cohort. **(B)** Enrichr visualization of biological process Gene Ontology (GO) terms. The most significant GO terms are represented in pink, Kyoto Encyclopedia of Genes and Genomes (KEGG) terms in grey, and genes are represented in green. For easier visualization, the nodes have been pooled into two categories: extracellular matrix organization and muscle contraction.

A mutation burden analysis across all known AIS-associated genes was conducted to assess for enrichment of rare variants in the affected cohort compared with unaffected cohort. The number of rare (MAF<0.1%), nonsynonymous variants in these population is presented in Supplementary Table 1 and rare nonsynonymous variants were identified in 10 known AIS-associated genes (*BNC2, KIF7, COL11A1/2, FBN1/2, HSPG2, POC5, ADGRG6,* and *FAT3*). *BNC2* and *KIF7* only had variants in affected individuals and variants in *BNC2*, the gene that carries the most significant risk polymorphisms, were all benign/likely benign.

The list of genetic variants that passed the filtering criteria of our analysis is presented in Table 2 for cases with solved or strong candidate variants, including categories 1−3, and Table 3 for cases with moderate candidate variants, category 4, respectively. In total, 137 candidate genes were identified, 116 of which were unique, 13 (*ATP7B, COL6A3, CUL7, DPP6, FAT4, FBN2, LAMA2, MYH7, NSD2, SBF1, SETD5, TRIO,* and *TRPS1*) were shared by two families, and 4 (*GJB2, HSPG2, MYH2*, and *RYR1*) were shared by three families. Candidate variants included coding and noncoding point mutations, along with structural variants, showing the utility of WGS. Candidate genes included previously AIS-associated genes (e.g. *CHD7, COL11A1/2, FBN1/2, HSPG2, KIF7*), as well as genes associated with other musculoskeletal and developmental syndromes where scoliosis is a known phenotype (e.g. *RYR1, GJB2, MYH2, MYH7*).

**Table 2.**
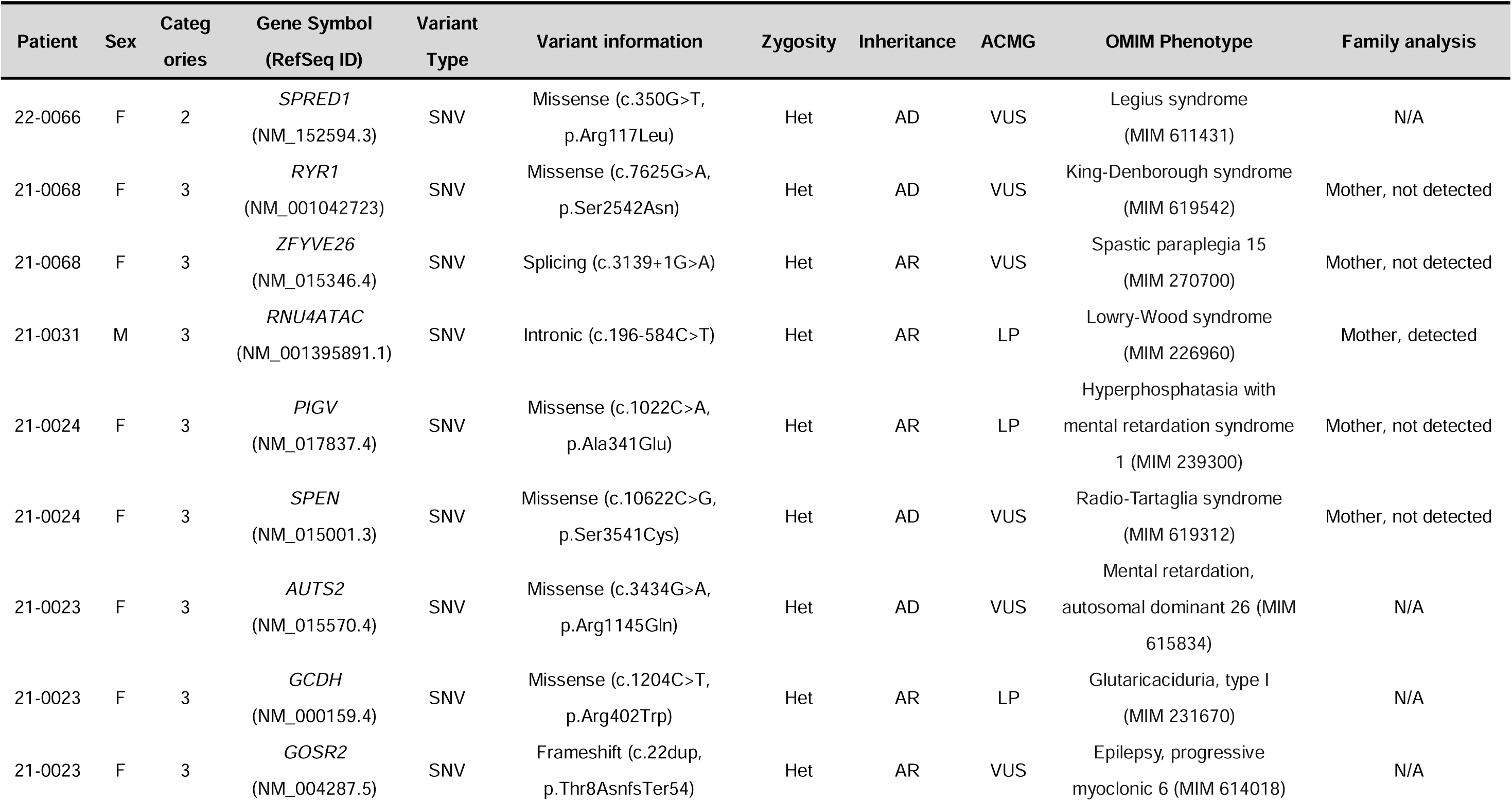
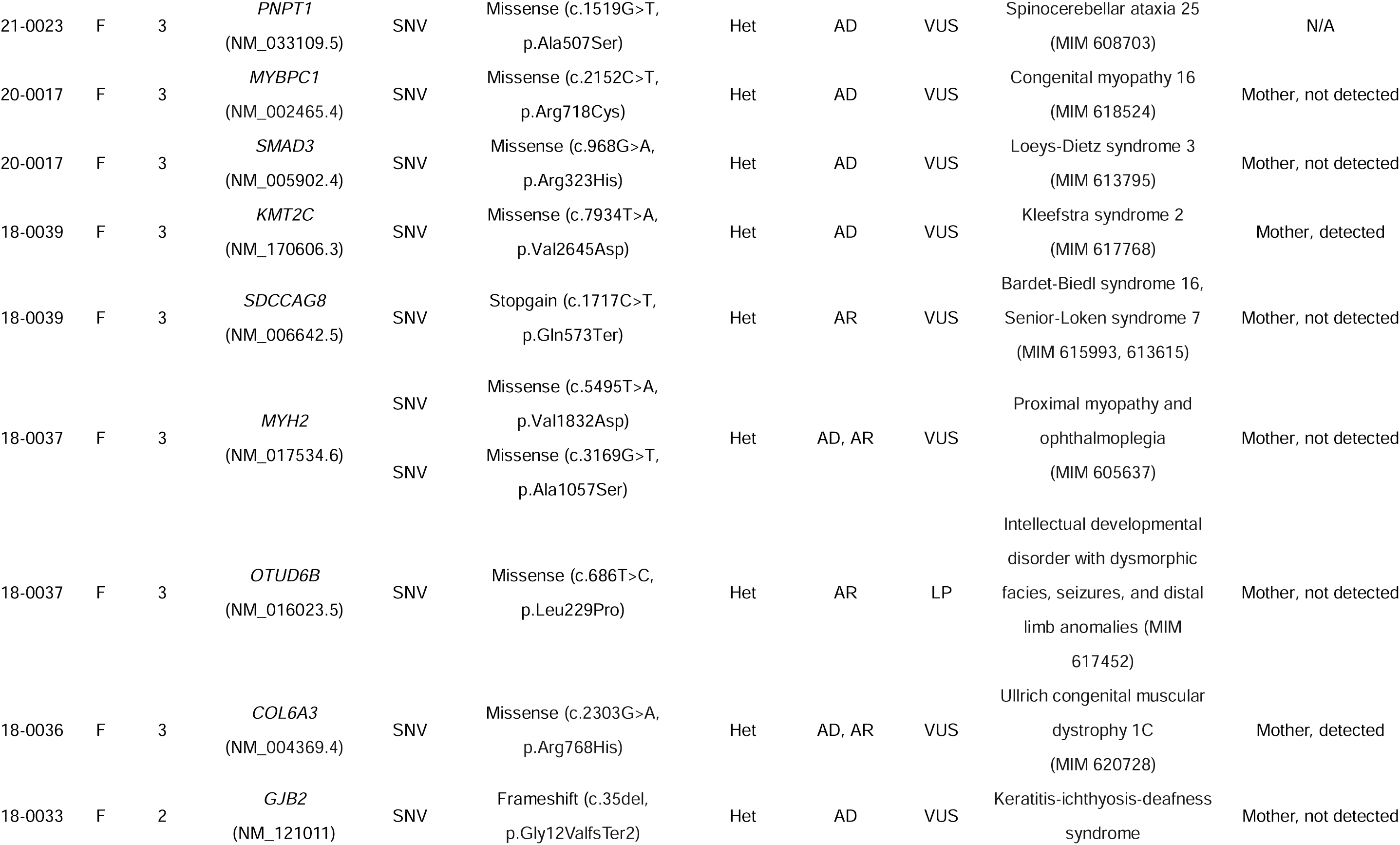
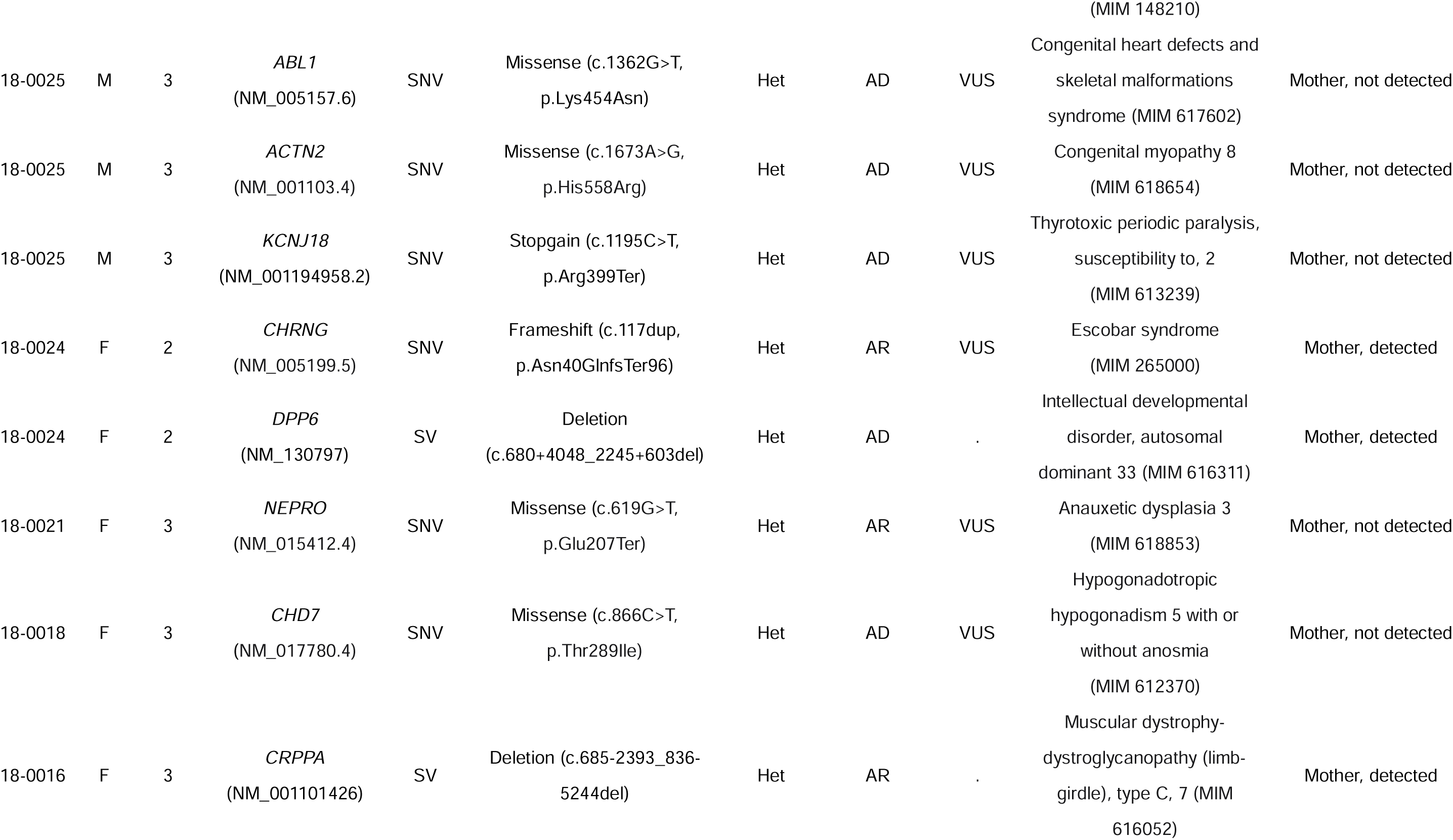
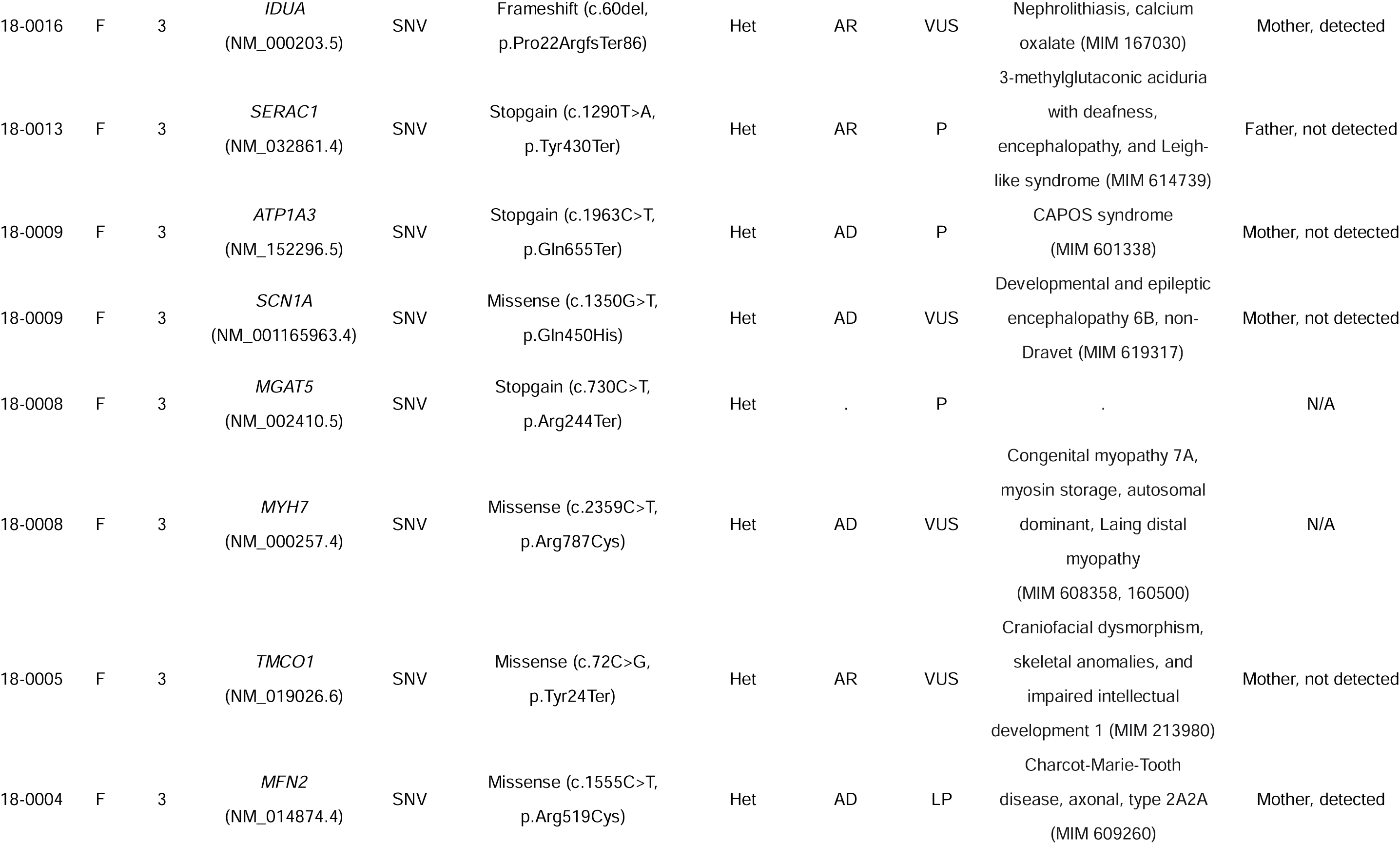
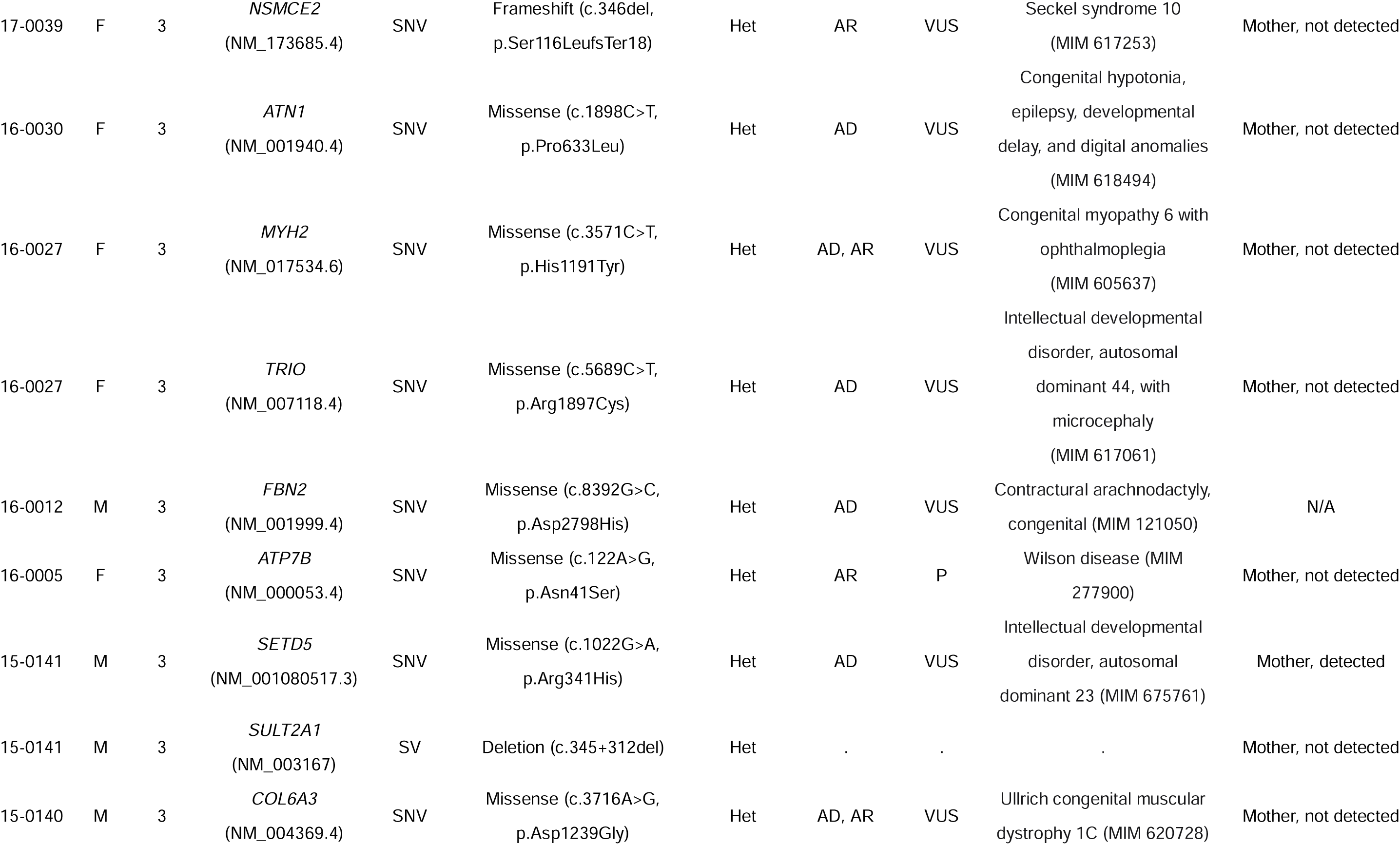
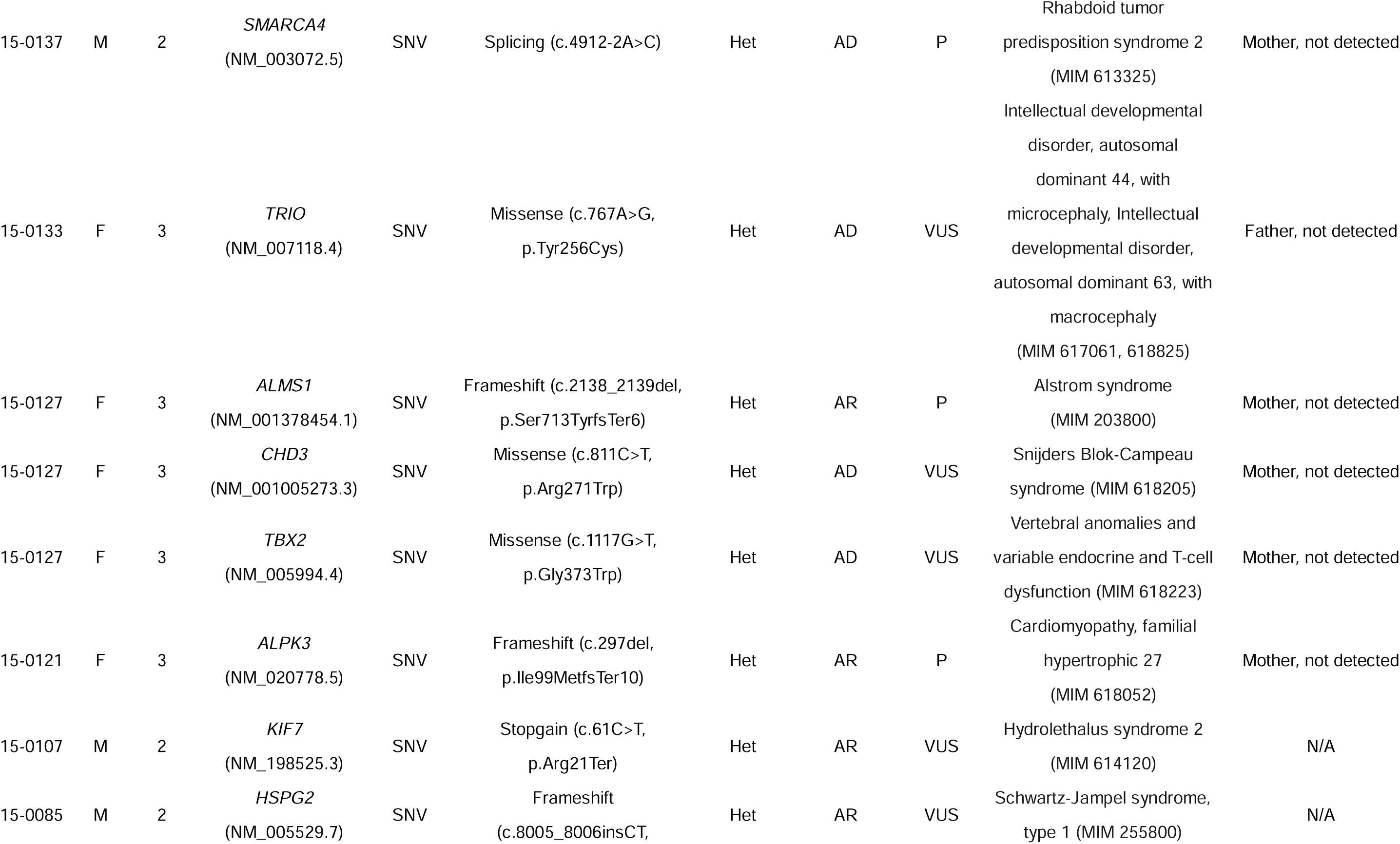
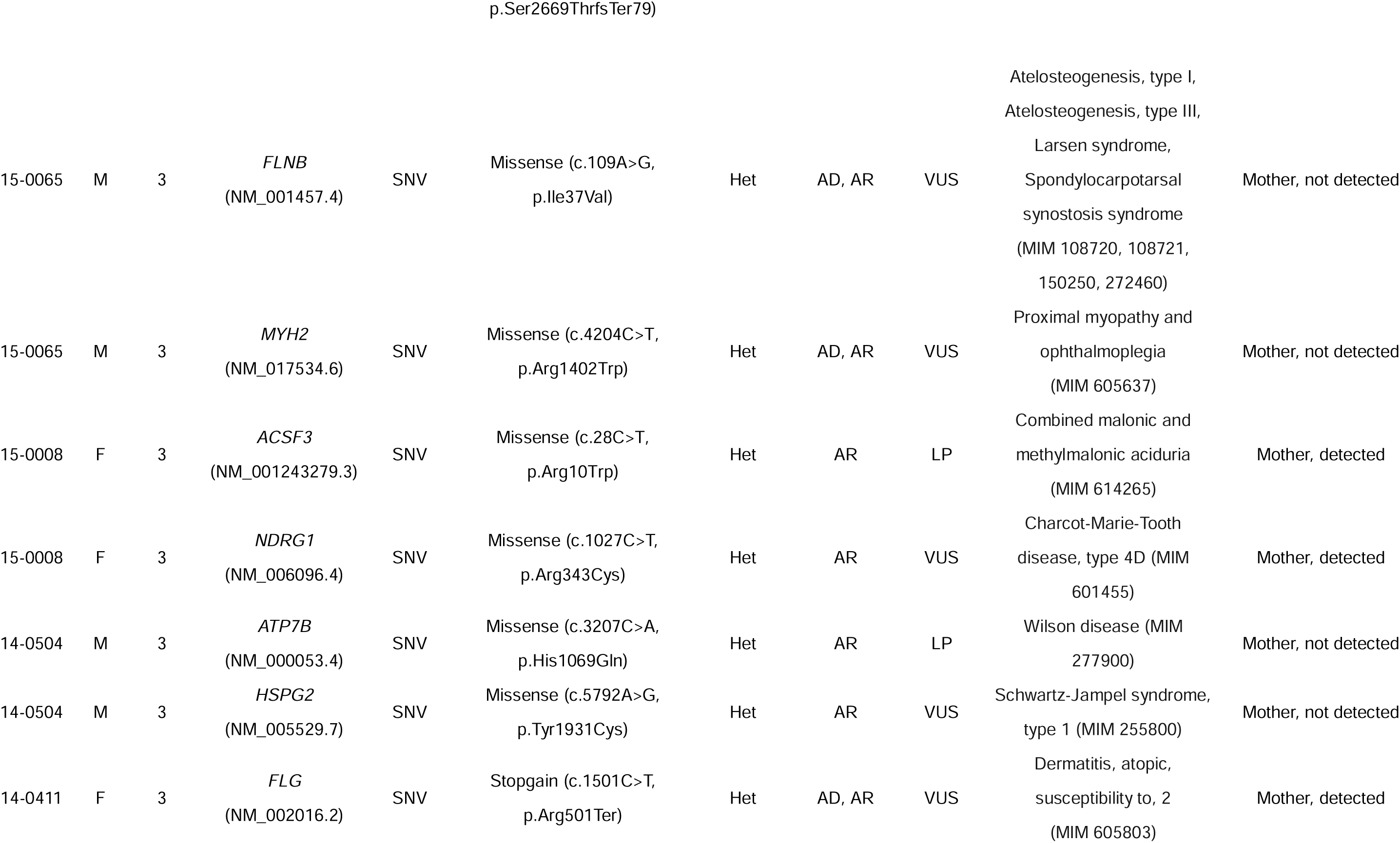
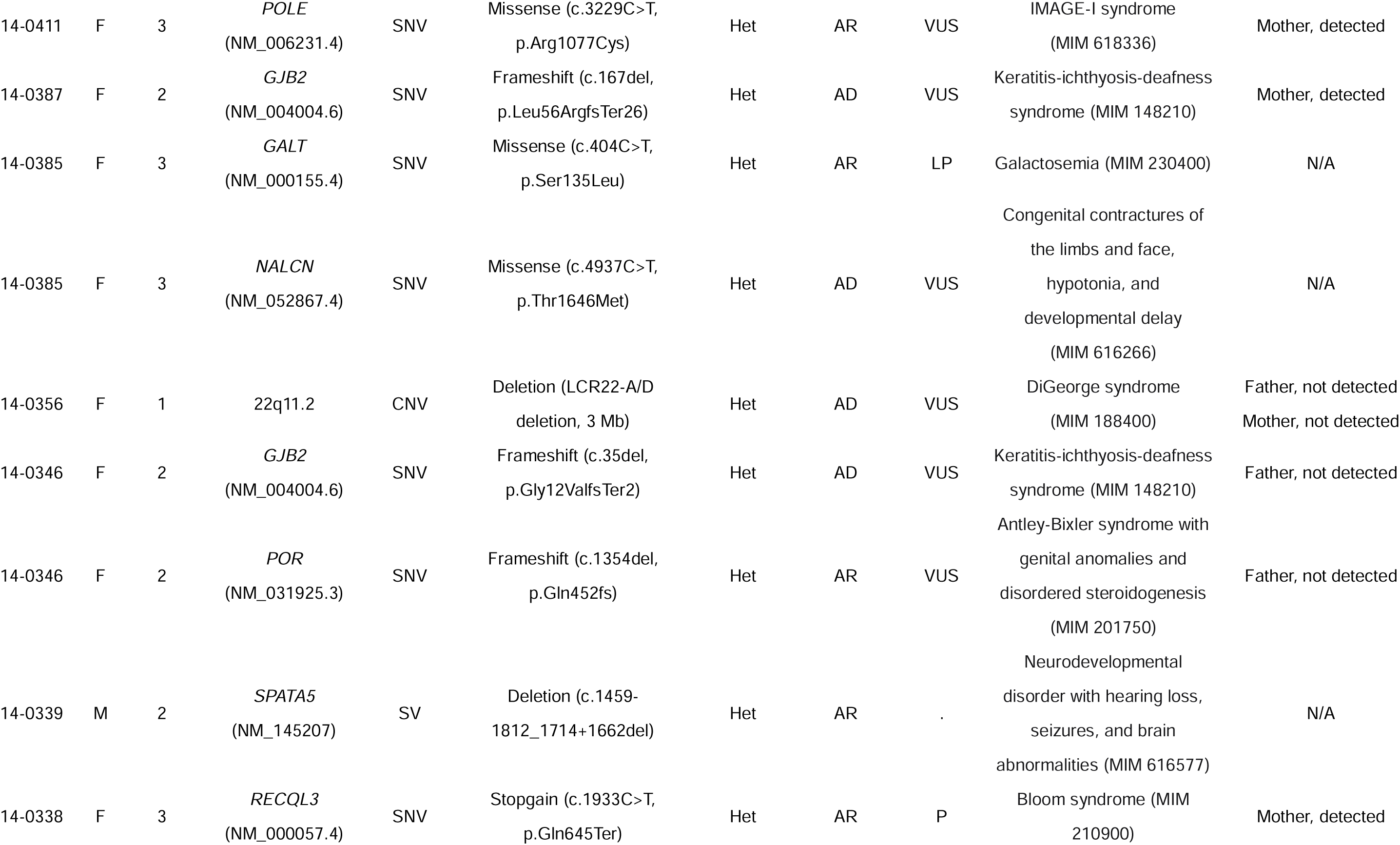
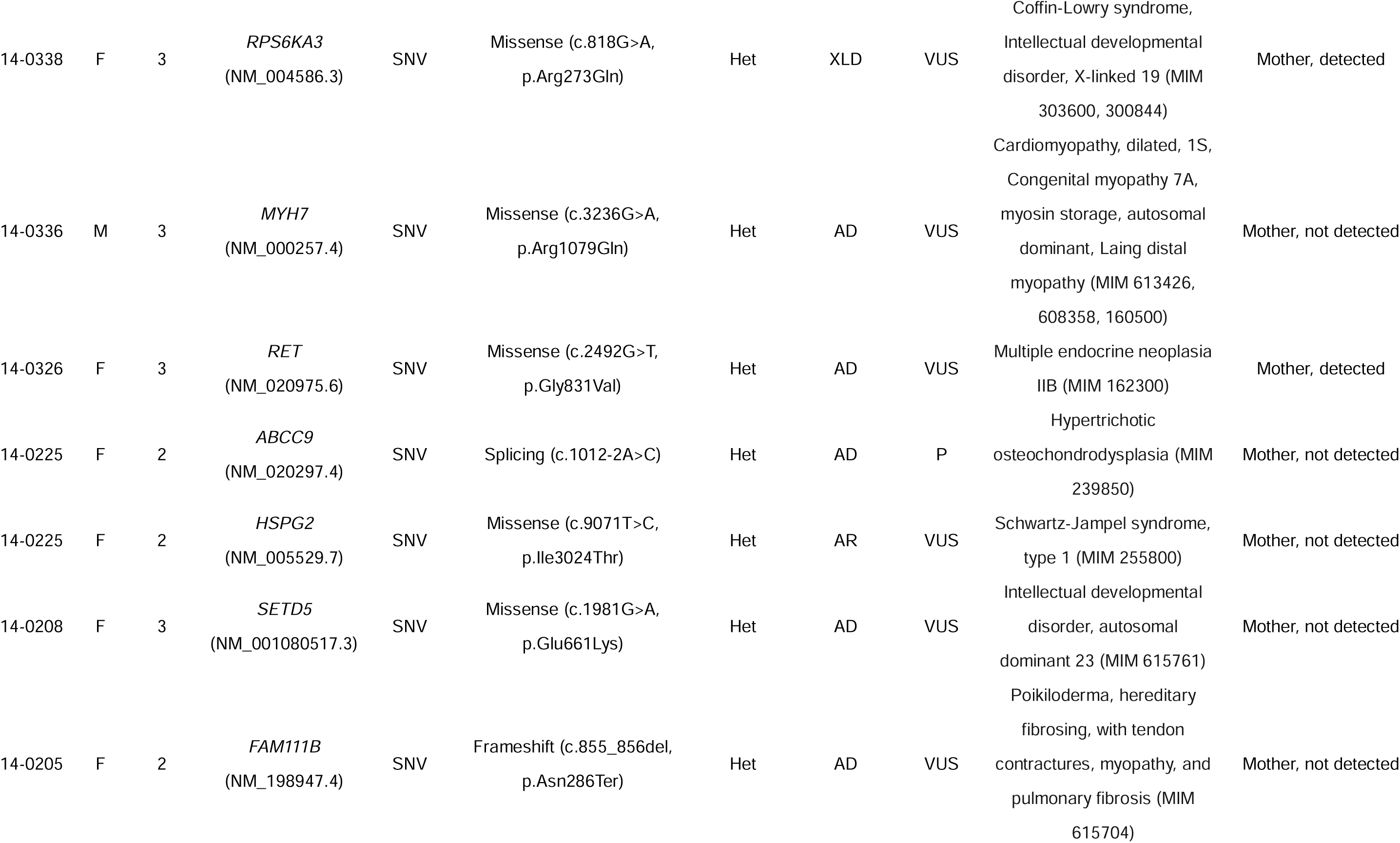
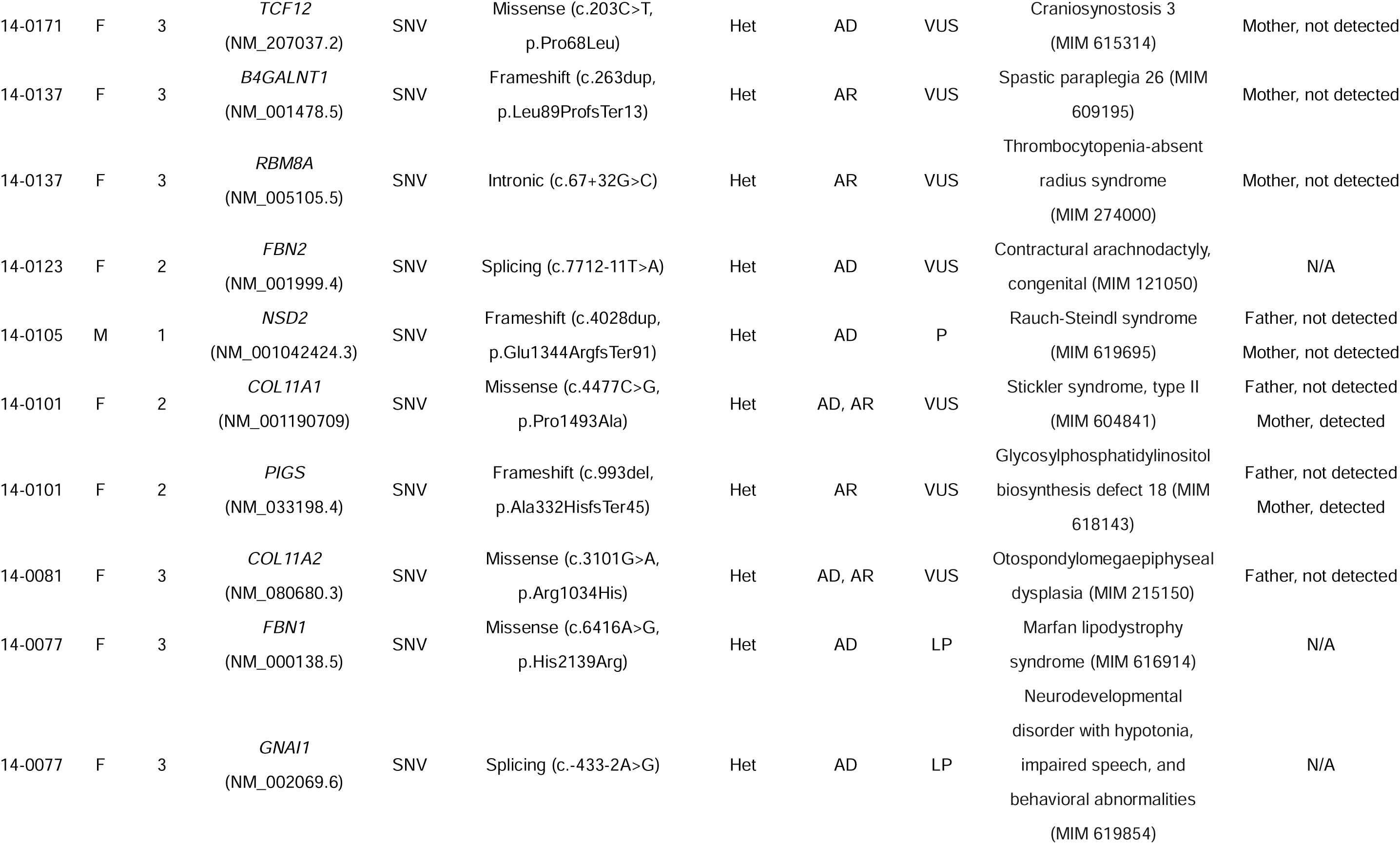
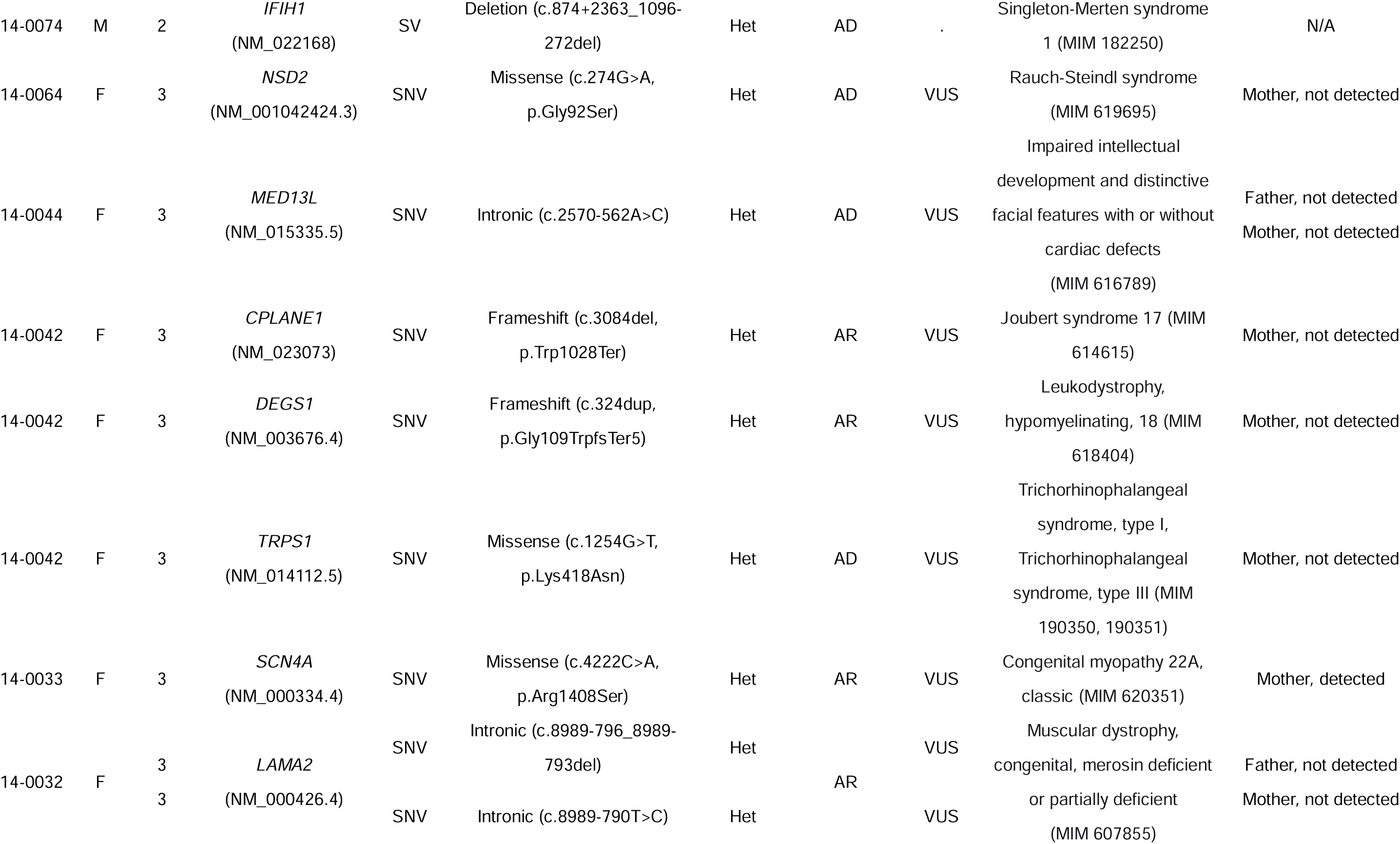
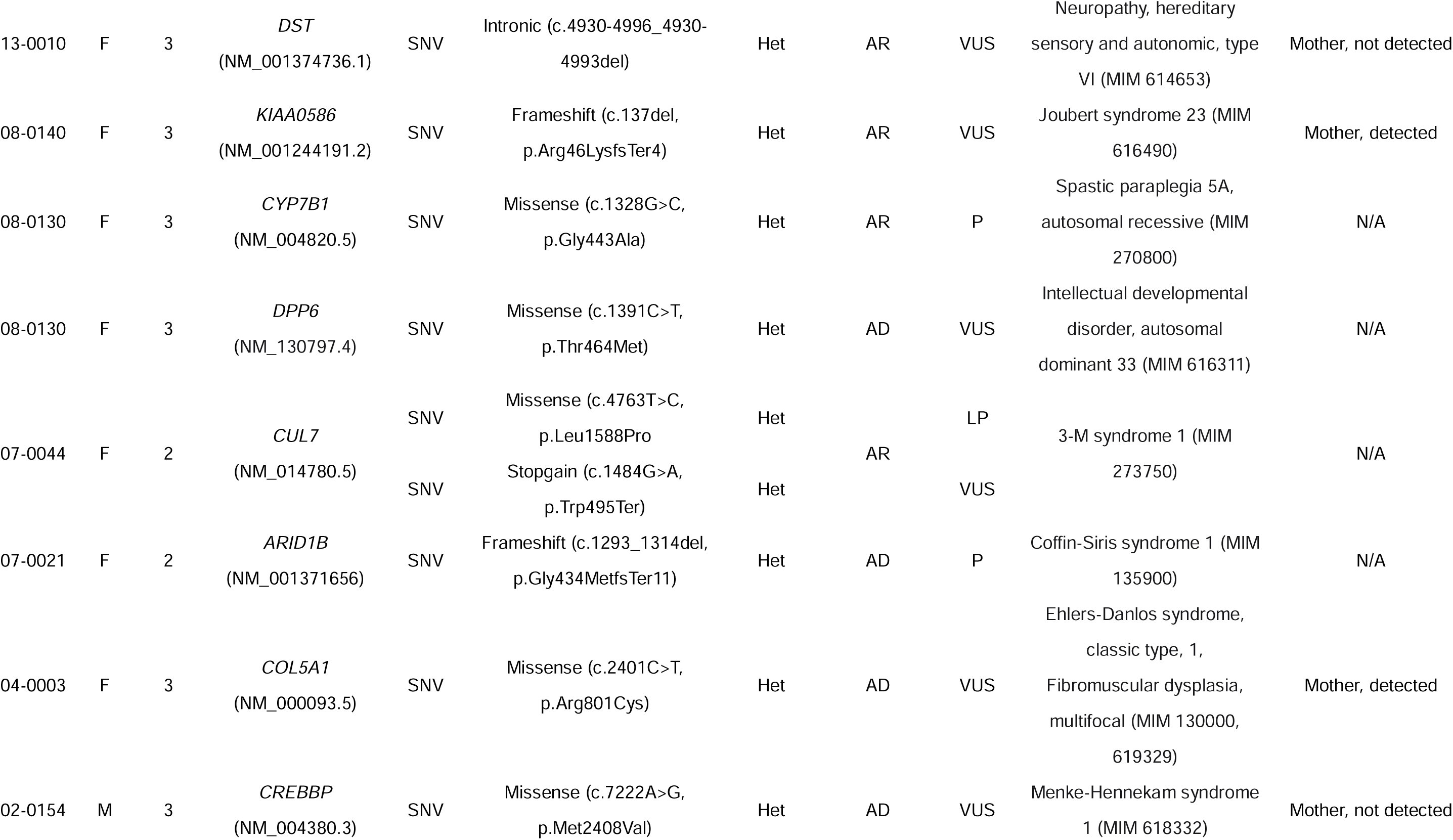
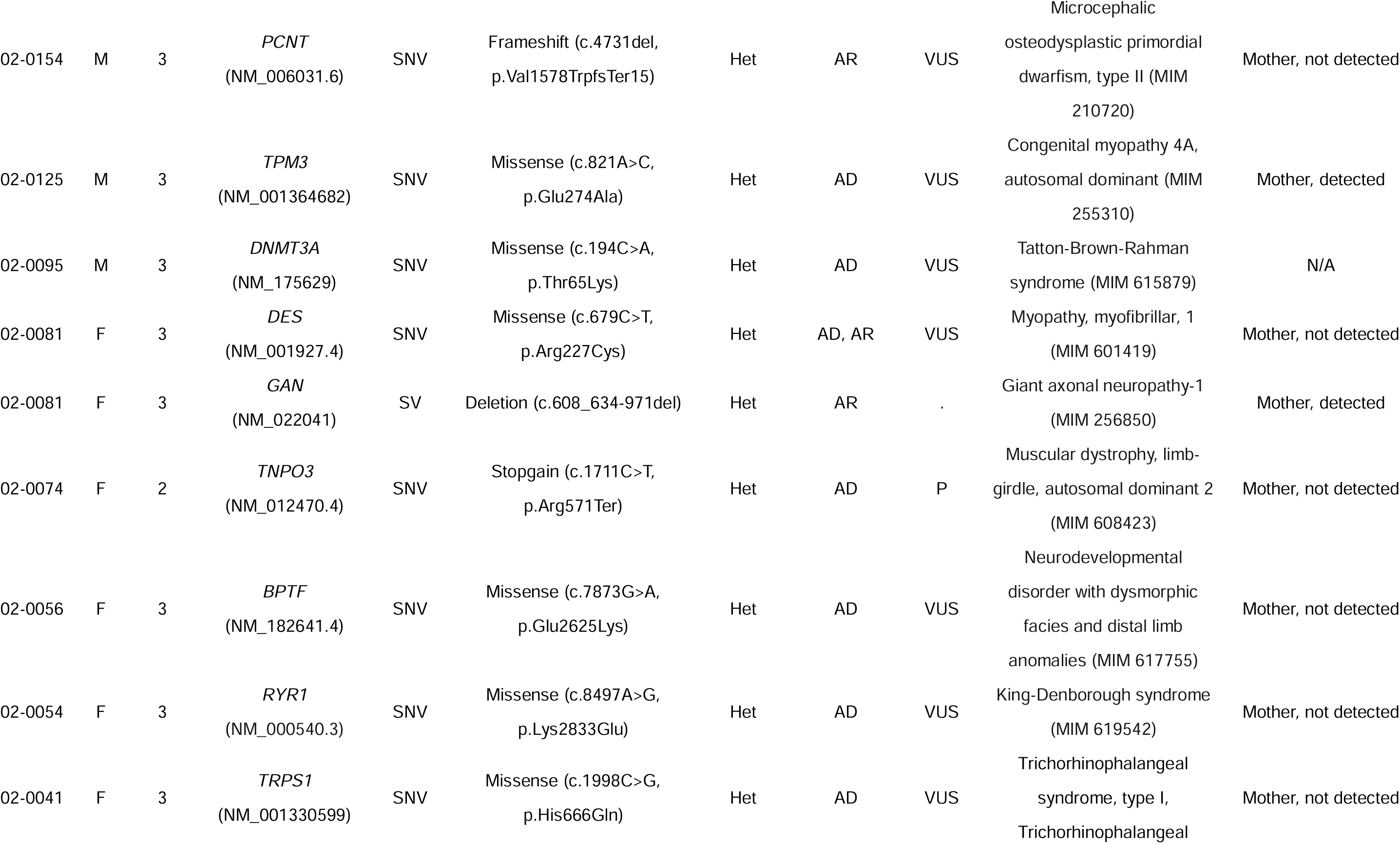
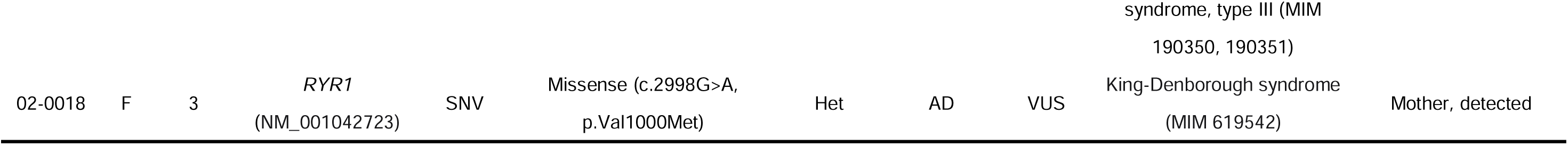
Cases with solved or strong candidate variants of adolescent idiopathic scoliosis (AIS)

**Table 3.**
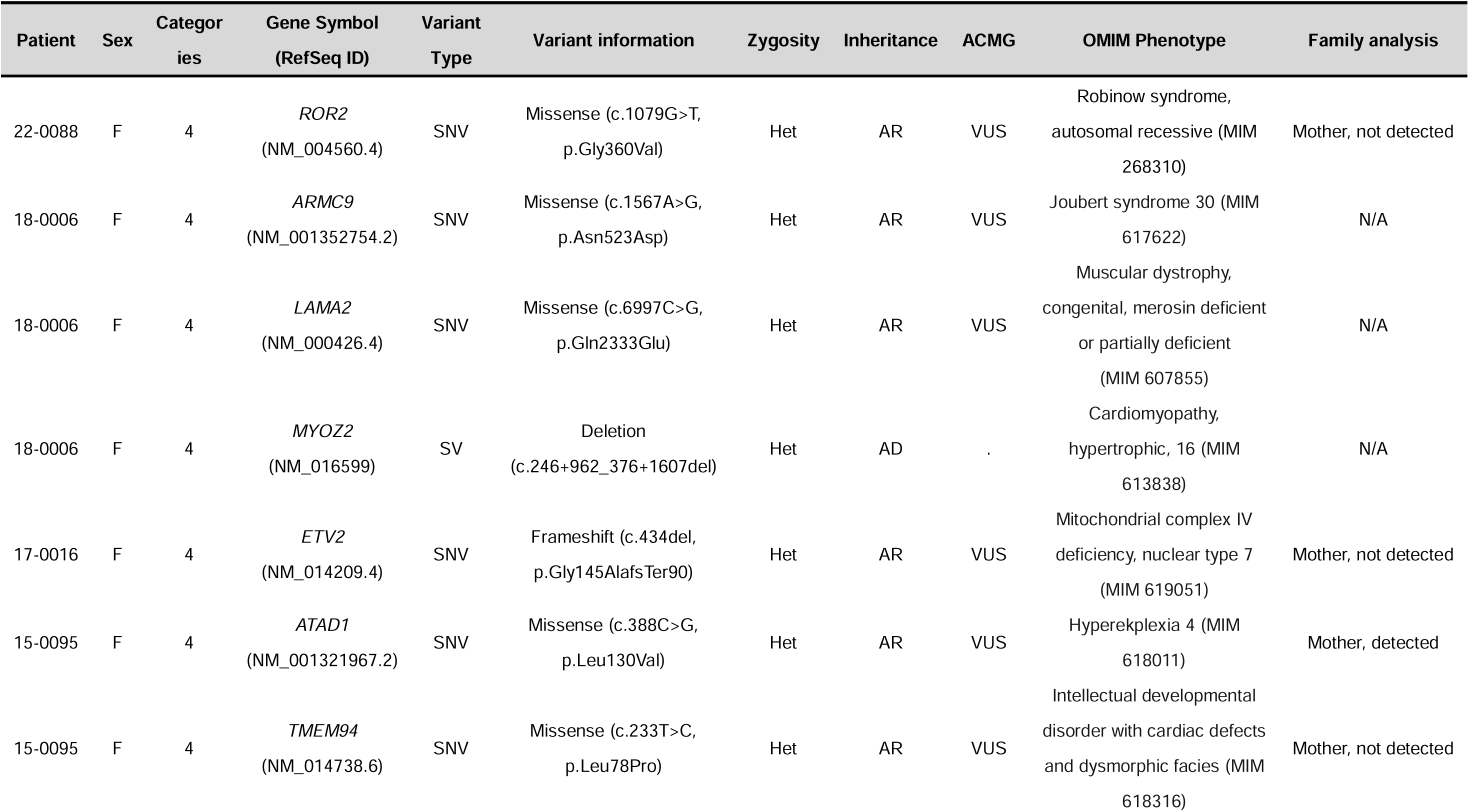
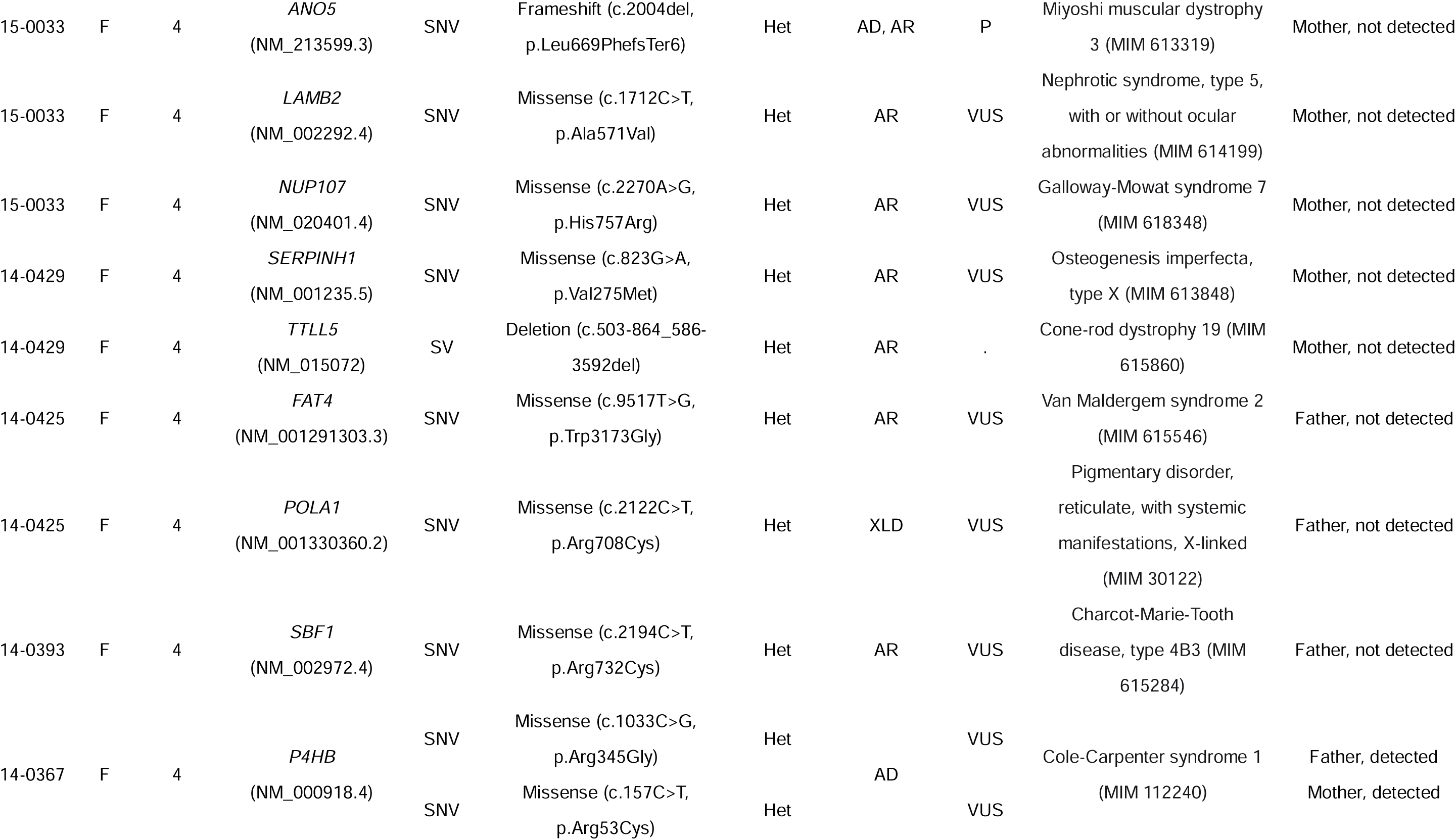
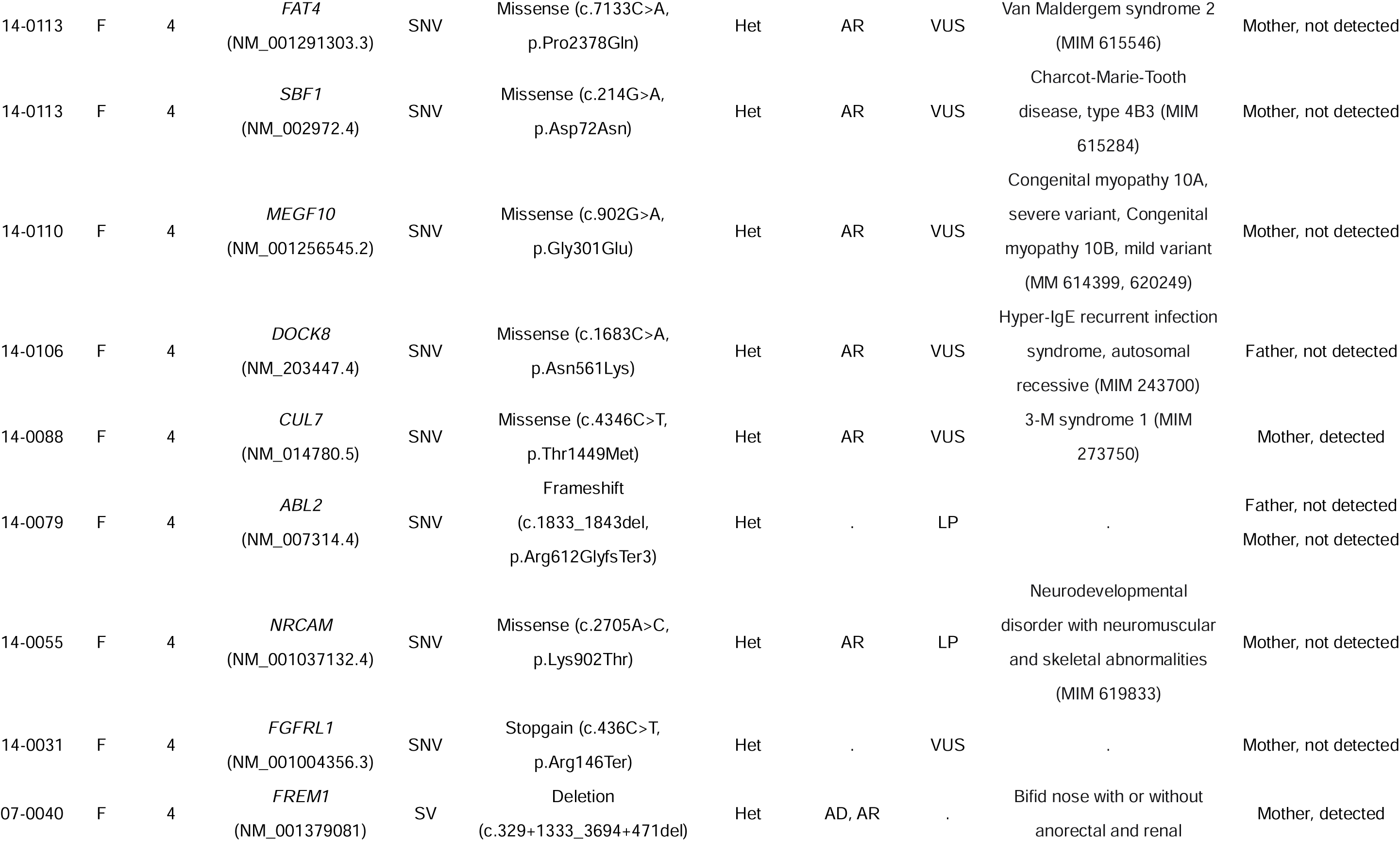
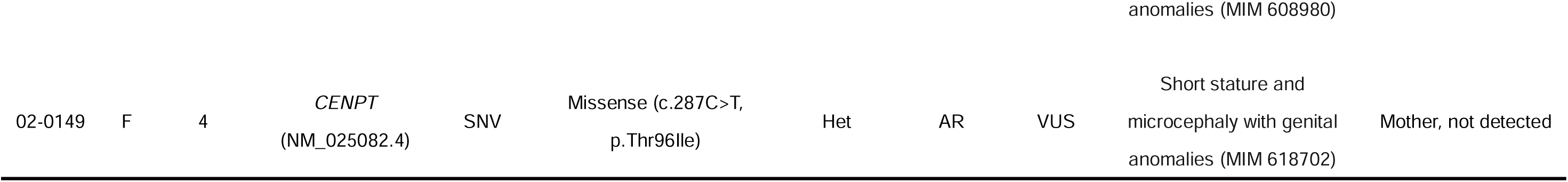
Cases with moderate candidate variants of adolescent idiopathic scoliosis (AIS)

### Gene Set Overrepresentation Analyses

To understand the connections between the candidate genes, we created an interaction map using the STRING database.^54^ We saw that 87 out of 137 candidate genes had at least one interactor (Supplementary Figure 1A). Furthermore, we identified 3 interconnected clusters that included the majority of the candidate genes (Supplementary Figure 1B). Cluster 1 included DNA-interacting proteins, including gene expression regulators like *ARID1B*, *CHD3*, *CREBBP*, and *KMT2C*. Cluster 2 included proteins involved in muscle contraction like *ACTN2*, *MYH2*, *MYH7*, and *SCN1A*. Cluster 3 included extracellular matrix (ECM) proteins like *COL11A1*, *FBN1*, *HSPG2*, and *LAMA2*.

To understand the biological functions of identified genes which were overrepresented with damaging variants within the AIS families, we performed an over-representation analysis based on the Mammalian Phenotype Ontology database of our resulting filtered gene lists. We entered the MAF <0.05 gene lists from individual patients into GO enrichment analysis to identify overrepresented gene ontologies: Cellular Component terms in Table 3 and Molecular Function terms in Table 4. The most overrepresented GO Cellular Component categories were “Z disc” (*p* = 9.21 × 10^-5^, 11.21 fold enrichment), “Endoplasmic reticulum lumen” (*p* = 5.17 × 10^-3^, 5.22 fold enrichment), “laminin-11 complex” (*p* = 6.87 × 10^-3^, >100 fold enrichment), and “protein complex involved in cell-matrix adhesion” (*p* = 7.67 × 10^-3^, 32.14 fold enrichment). The most overrepresented GO Molecular Function categories were “ATP-dependent activity” (*p* = 3.00 × 10^-3^, 4.34 fold enrichment), “extracellular matrix constituent conferring elasticity” (*p* = 6.36 × 10^-3^, 54.64 fold enrichment), “ATP binding” (*p* = 6.46 × 10^-3^, 2.68 fold enrichment), and “integrin binding” (*p* = 9.24 × 10^-3^, 7.97 fold enrichment). Figure 2B displays a visual representation of Enrichr and the first ten clusters in terms of *p*-value were linked to binding activities. The top enriched GO Biological Process term was “extracellular matrix organization” (*p* = 1.12 × 10^-8^) and “muscle contraction” (p = 2.28 × 10^-8^). Furthermore, the top enriched KEGG term was “ECM-receptor interaction” (*p* = 3.02 × 10^-4^).

**Table 4.** Overrepresented Gene Ontology, Cellular Component terms using the combined Minor Allele Frequency (MAF) < 0.05 gene list from all probands.

| Term | Count | % | P-Value | Genes | Fold Enrichment |
| --- | --- | --- | --- | --- | --- |
| laminin-11 complex | 2 | 0.02 | $6.87 \times 10^{-3}$ | *LAMB2, LAMC1* | > 100 |
| collagen type XI trimer | 2 | 0.02 | $1.27 \times 10^{-2}$ | *COL11A2, COL11A1* | 91.06 |
| protein complex involved in cell-matrix adhesion | 3 | 0.09 | $7.67 \times 10^{-3}$ | *LAMA2, LAMB2, LAMC1* | 32.14 |
| myosin filament | 3 | 0.13 | $1.93 \times 10^{-2}$ | *MYH7, MYBPC1, MYH2* | 22.77 |
| Z disc | 8 | 0.71 | $9.21 \times 10^{-5}$ | *MYH7, DES, RYR1, FLNB, ACTN2, DST, SCN1A, MYOZ2* | 11.21 |
| nuclear matrix | 5 | 0.70 | $4.03 \times 10^{-2}$ | *RECQL3, DNMT3A, ATN1, SMARCA4, POLA1* | 7.11 |
| endoplasmic reticulum lumen | 9 | 1.72 | $5.17 \times 10^{-3}$ | *COL5A1, SERPINH1, COL6A3, P4HB, COL11A2, LAMB2, LAMC1, FBN1, COL11A1* | 5.22 |
| monoatomic ion channel complex | 8 | 1.90 | $3.83 \times 10^{-2}$ | *ABCC9, NALCN, RYR1, SCN4A, CHRNG, DPP6, KCNJ18, SCN1A* | 4.21 |
| actin cytoskeleton | 10 | 2.84 | $3.42 \times 10^{-2}$ | *TPM3, MYH7, AUTS2, ABL1, FLNB, ACTN2, ABL2, DST, MYOZ2, MYH2* | 3.52 |

### Allelic Test of Association for SNPs

We attempted to compare SNPs between our patient cohort and a general population cohort of the similar ethnic background to determine their association with AIS disease. Pérez-Machado et al.^7^ proposed SNPs with the most statistically significant associations with AIS using 10 previous studies and 1387 published associations reported up to 2019. We performed allelic tests of association in our patient cohort for the 15 SNPs proposed by Pérez-Machado et al. Table 5 lists the genes and SNPs of these results and showed a significant association of genetic variants rs12946942, located between *SOX9* and *KCNJ2*, with AIS. A significant association of loci (rs10756785 and rs3904778) of *BNC2* gene and loci (rs7633294) of *MAGI1* with AIS were also found.

**Table 5.**
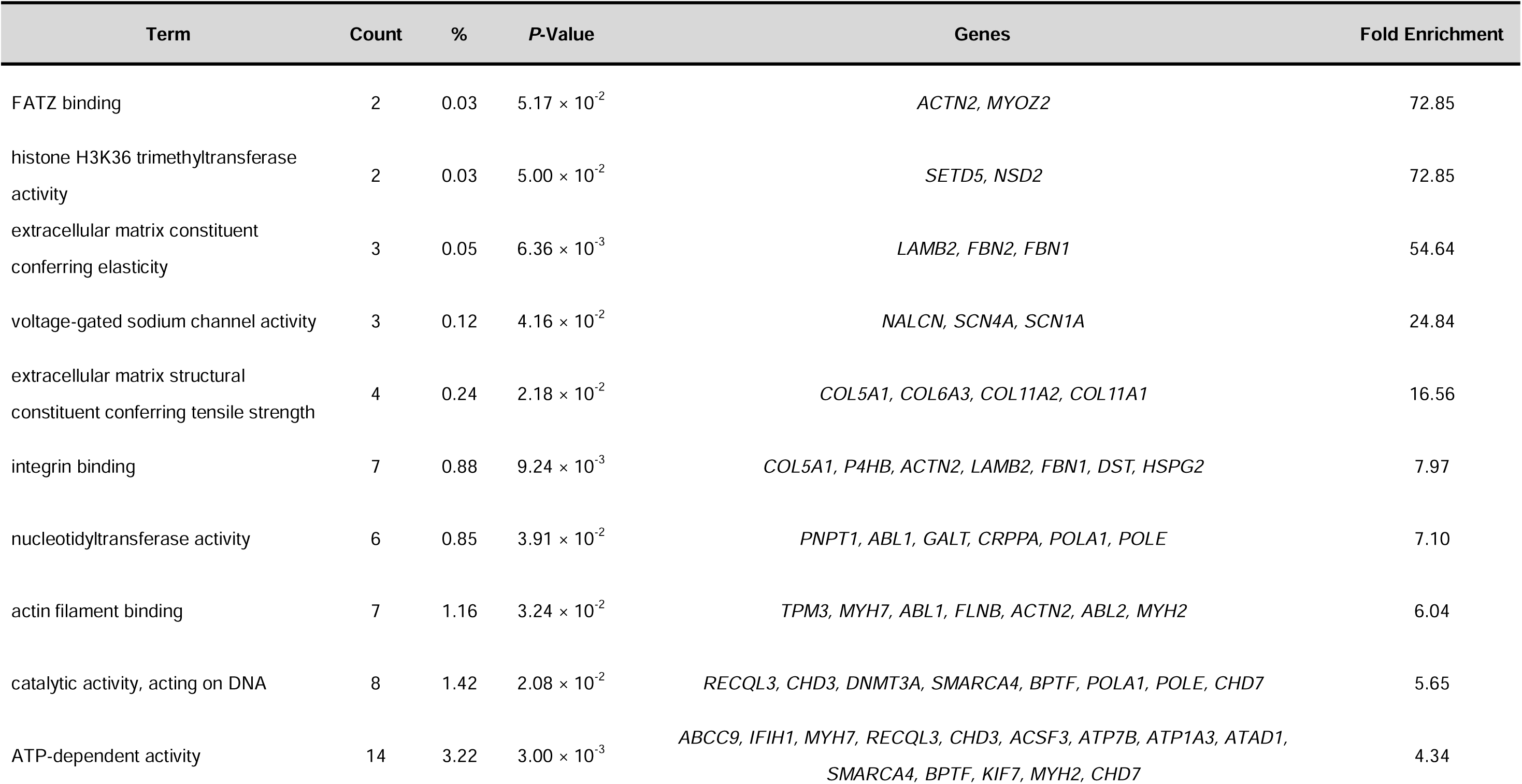
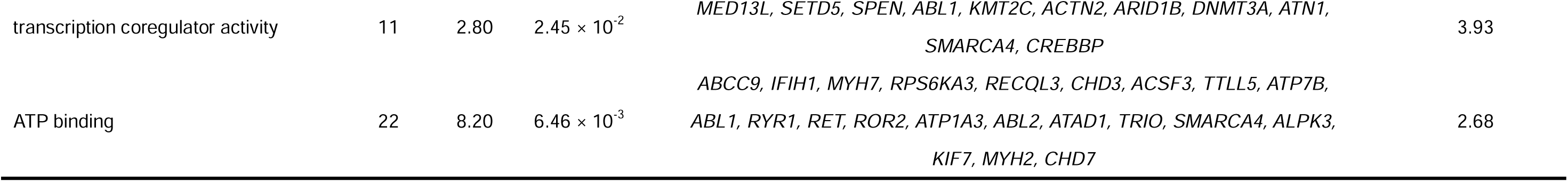
Overrepresented Gene Ontology, Molecular Function terms using the combined Minor Allele Frequency (MAF) 1 < 0.05 gene list from all probands.

| Term | Count | % | P-Value | Genes | Fold Enrichment |
| --- | --- | --- | --- | --- | --- |
| FATZ binding | 2 | 0.03 | $5.17 \times 10^{-2}$ | *ACTN2, MYOZ2* | 72.85 |
| histone H3K36 trimethyltransferase activity | 2 | 0.03 | $5.00 \times 10^{-2}$ | *SETD5, NSD2* | 72.85 |
| extracellular matrix constituent conferring elasticity | 3 | 0.05 | $6.36 \times 10^{-3}$ | *LAMB2, FBN2, FBN1* | 54.64 |
| voltage-gated sodium channel activity | 3 | 0.12 | $4.16 \times 10^{-2}$ | *NALCN, SCN4A, SCN1A* | 24.84 |
| extracellular matrix structural constituent conferring tensile strength | 4 | 0.24 | $2.18 \times 10^{-2}$ | *COL5A1, COL6A3, COL11A2, COL11A1* | 16.56 |
| integrin binding | 7 | 0.88 | $9.24 \times 10^{-3}$ | *COL5A1, P4HB, ACTN2, LAMB2, FBN1, DST, HSPG2* | 7.97 |
| nucleotidyltransferase activity | 6 | 0.85 | $3.91 \times 10^{-2}$ | *PNPT1, ABL1, GALT, CRPPA, POLA1, POLE* | 7.10 |
| actin filament binding | 7 | 1.16 | $3.24 \times 10^{-2}$ | *TPM3, MYH7, ABL1, FLNB, ACTN2, ABL2, MYH2* | 6.04 |
| catalytic activity, acting on DNA | 8 | 1.42 | $2.08 \times 10^{-2}$ | *RECQL3, CHD3, DNMT3A, SMARCA4, BPTF, POLA1, POLE, CHD7* | 5.65 |
| ATP-dependent activity | 14 | 3.22 | $3.00 \times 10^{-3}$ | *ABCC9, IFIH1, MYH7, RECQL3, CHD3, ACSF3, ATP7B, ATP1A3, ATAD1, SMARCA4, BPTF, KIF7, MYH2, CHD7* | 4.34 |
| transcription coregulator activity | 11 | 2.80 | $2.45 \times 10^{-2}$ | *MED13L, SETD5, SPEN, ABL1, KMT2C, ACTN2, ARID1B, DNMT3A, ATN1, SMARCA4, CREBBP* | 3.93 |
| ATP binding | 22 | 8.20 | $6.46 \times 10^{-3}$ | *ABCC9, IFIH1, MYH7, RPS6KA3, RECQL3, CHD3, ACSF3, TTLL5, ATP7B, ABL1, RYR1, RET, ROR2, ATP1A3, ABL2, ATAD1, TRIO, SMARCA4, ALPK3, KIF7, MYH2, CHD7* | 2.68 |

**Table 6.** Allelic test of association for SNPs previously associated with adolescent idiopathic scoliosis (AIS)

| SNP_ID | Genes | Context | SNP | Affected |  |  | 1000G Controls |  |  | CHIS<br>Q | p-Value | OR [95% CI] |
| --- | --- | --- | --- | --- | --- | --- | --- | --- | --- | --- | --- | --- |
|  |  |  |  | Ref | Alt | Alt_Fq | Ref | Alt | Alt_Fq |  |  |  |
| rs12946942 | SOX9, KCNJ2 | Intergenic | chr17:71240857:G>T | 159 | 7 | 0.042 | 1933 | 313 | 0.139 | 12.69 | $3.68 \times 10^{-4}$ | 0.27 [0.13–0.59] |
| rs10756785 | BNC2 | Intron | chr9:16700839:C>T | 31 | 135 | 0.813 | 699 | 1547 | 0.689 | 11.35 | $7.56 \times 10^{-4}$ | 1.97 [2.94–1.32] |
| rs3904778 | BNC2 | Intron | chr9:16681995:C>G | 30 | 136 | 0.819 | 663 | 1583 | 0.705 | 9.891 | $1.66 \times 10^{-3}$ | 1.90 [2.85–1.27] |
| rs7633294 | MAGI1 | Intron | chr3:65650810:A>G | 42 | 124 | 0.747 | 834 | 1412 | 0.629 | 9.356 | $2.22 \times 10^{-3}$ | 1.74 [2.50–1.22] |
| rs4513093 | CDH13 | Intron | chr16:82872937:A>G | 17 | 149 | 0.898 | 343 | 1903 | 0.847 | 3.081 | $7.92 \times 10^{-2}$ | 1.58 [2.64–0.94] |
| rs7593846 | MEIS1 | Intron | chr2:66664256:T>C | 139 | 27 | 0.163 | 1777 | 469 | 0.209 | 2.017 | $1.56 \times 10^{-1}$ | 0.74 [0.48–1.13] |
| rs6137473 | FOXA2, PAX1 | Intergenic | chr20:21904055:G>A | 87 | 79 | 0.476 | 1074 | 1172 | 0.522 | 1.305 | $2.53 \times 10^{-1}$ | 0.83 [1.14–0.61] |
| rs6047663 | PAX1 | Regulatory region | chr20:21841272:C>T | 83 | 83 | 0.500 | 1047 | 1199 | 0.534 | 0.711 | $4.00 \times 10^{-1}$ | 0.87 [1.20–0.64] |
| rs13398147 | PAX3/EPHA4 | Intergenic | chr2:221895559:C>T | 125 | 41 | 0.247 | 1754 | 492 | 0.219 | 0.701 | $4.03 \times 10^{-1}$ | 1.17 [0.81–1.69] |
| rs678741 | LBX1, AS1 | Intron | chr10:101237824:G>A | 60 | 106 | 0.639 | 884 | 1362 | 0.606 | 0.670 | $4.13 \times 10^{-1}$ | 1.15 [1.59–0.83] |
| rs11190870 | LBX1 | Intergenic | chr10:101219450:T>C | 110 | 56 | 0.337 | 1418 | 828 | 0.369 | 0.653 | $4.19 \times 10^{-1}$ | 0.87 [0.63–1.22] |
| rs6570507 | GPR126 | Intron | chr6:142358435:G>A | 107 | 59 | 0.355 | 1464 | 782 | 0.348 | 0.036 | $8.50 \times 10^{-1}$ | 1.03 [0.74–1.44] |
| rs241215 | AJAP1 | Intergenic | chr1:4544044:T>A | 73 | 93 | 0.560 | 971 | 1275 | 0.568 | 0.035 | $8.52 \times 10^{-1}$ | 0.97 [1.33–0.71] |
| rs4940576 | BCL2 | Intron | chr18:63181406:T>C | 43 | 123 | 0.741 | 576 | 1670 | 0.744 | 0.005 | $9.42 \times 10^{-1}$ | 0.99 [1.41–0.69] |
| rs169311 | PAX1 | Intergenic | chr20:21981695:A>C | 89 | 77 | 0.464 | 1209 | 1037 | 0.462 | 0.003 | $9.57 \times 10^{-1}$ | 1.01 [0.74–1.38] |

### Representative Cases

The genetic variant information of representative cases in which we were able to identify the causative gene of AIS through WGS are presented. Patient 14-0356 was a trio with unaffected parents and was identified with a ∼3.0 Mbp deletion encompassing four low copy repeats (LCR) A to D in the chromosome 22q11.2 region (Figure 3). Comparison of coverage of the 22q11 region in this family showed that this variant was a *de novo* deletion occurred only in the proband. 22q11.2 proximal deletions are associated with DiGeorge syndrome (MIM 188400), which presents with several physiological and anatomical phenotypes, including scoliosis.^55^ The *TBX1* gene is located at region 22q11.2, and an haploinsufficiency of *TBX1* has been identified as the driver of for most of the physical malformations in patients with DiGeorge syndrome.^56^

**Figure 3.**
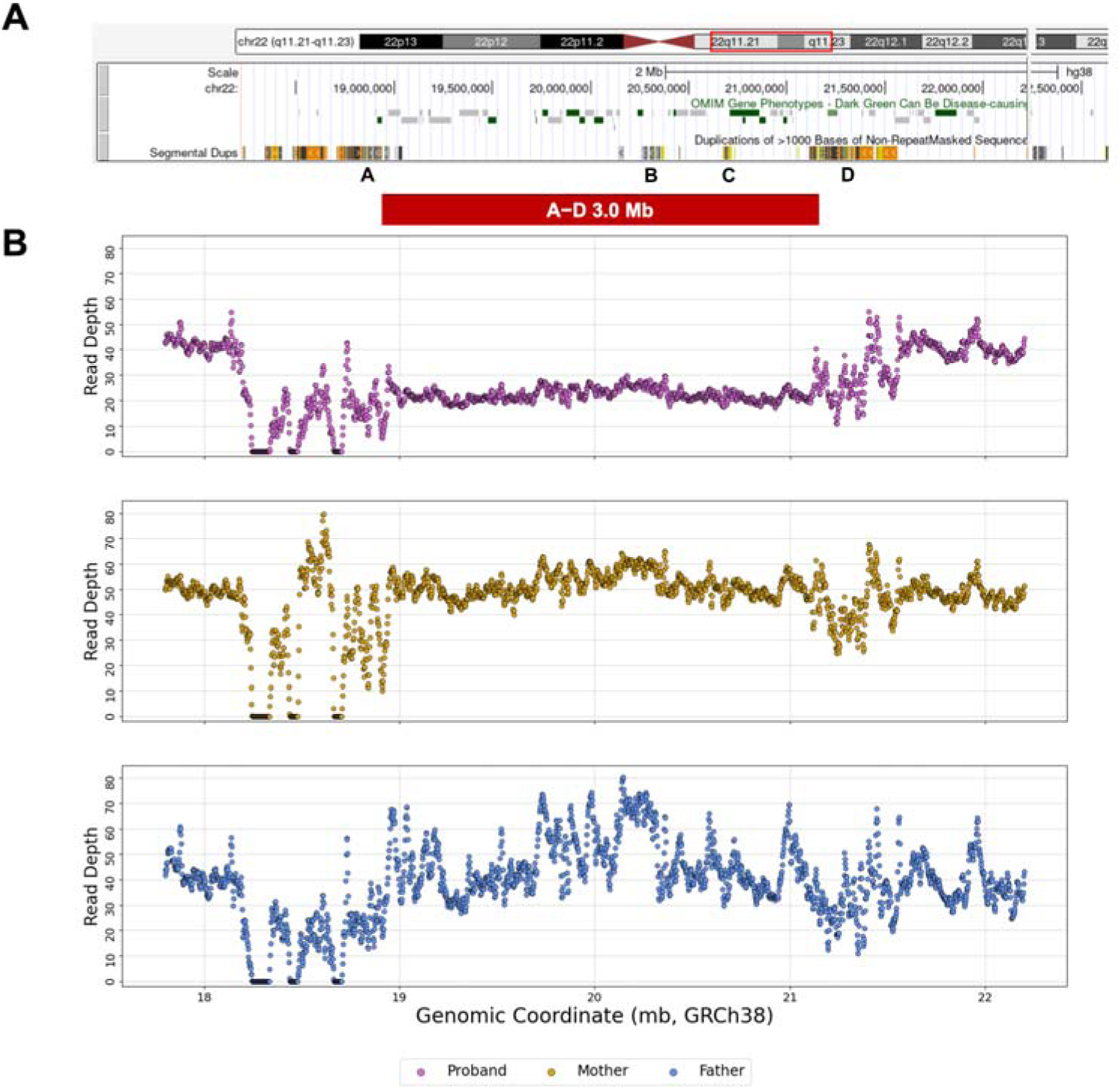
Genetic results of patient 14-0356. **(A)** Schematic overview of the chromosome 22q11.2 region from the UCSC Genome Browser. The region of the 22q11.2 chromosomal deletion in this patient spans approximately 3 Mb and contains four low copy repeats (LCR A-LCR D) distributed across this region. **(B)** Coverage of chromosome 22q11.2 region in this trio family (proband in purple, mother in yellow, and father in blue). Y-axis shows read depth and X-axis shows position in Kbs. *De novo* deletion region in proband is shown with red lines.

Patient 14-0105 was also a trio with unaffected parents and the genetic results can be seen in Figure 4. A frameshift variant (p.Glu1344ArgfsTer91) in the *NSD2* gene on chromosome 4 (chr4:1978831:G>GC) was identified with WGS. This variant was not found in the patient’s parents, indicating that it was a *de novo* event. Haploinsufficiency due to *de novo* heterozygous frameshift and missense variants in *NSD2* has been reported to cause a diverse spectrum of skeletal abnormalities, including scoliosis.^57^

**Figure 4.**
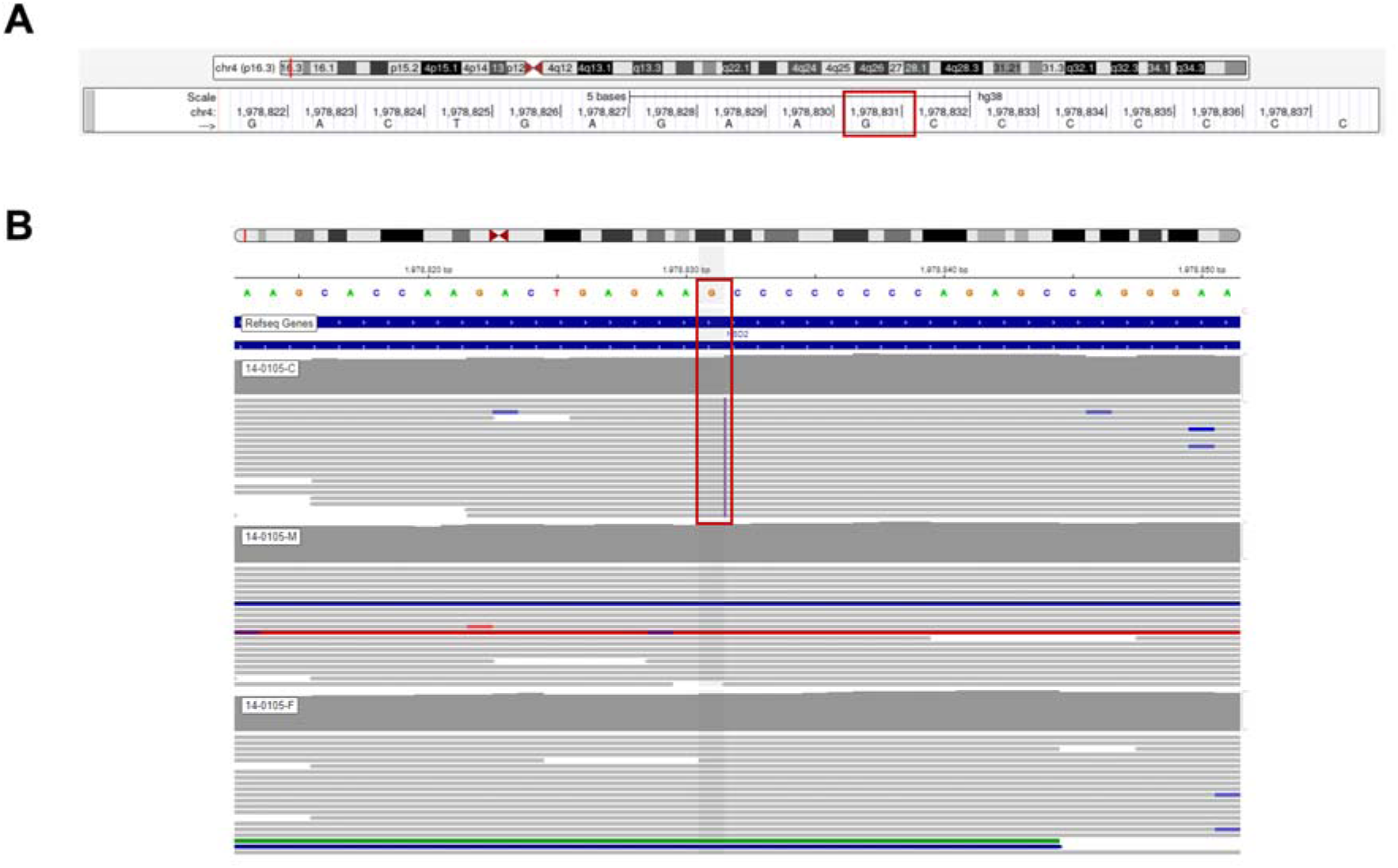
Genetic results of patient 14-0105. **(A)** Schematic figure from the UCSC Genome Browser. Note the mutation region in the *NSD2* gene on chromosome 4 in this patient. **(B)** Integrative Genomics Viewer image of trios (proband-mother-father). Note the *de novo* event, chr4:1,978,831:G>GC, results in a frameshift variant in *NSD2* (p.Glu1344ArgfsTer91).

## DISCUSSION

In the present study, we investigated the genetic landscape in 103 AIS probands utilizing WGS. To the best of our knowledge, this is the largest study to explore the comprehensive genomic landscape of an AIS cohort using WGS.

The lack of overlap in specific genes between different families suggests that AIS is a highly polygenic disorder involving variants in multiple genes, potentially suggesting that a combination of genetic/epigenetic and environmental factors are involved in the expression of different phenotypes. GO enrichment analysis revealed that the genes identified in this AIS cohort are mainly involved in “extracellular matrix organization” and “muscle contraction”. Allelic tests of association for SNPs showed that SNP risk alleles located between *SOX9* and *KCNJ2*, *BNC2*, and *MACl1* genes were significantly associated with AIS. A majority of the candidate genes in our list were associated with other complex syndromes with severe phenotypes, including but not limited to scoliosis. This could indicate that patients diagnosed with AIS may be presenting with milder cases of more severe syndromes.

Genes related to ECM organization were the first enriched category in our dataset. The ECM is a complex and dynamic component of all tissues that is made up of components including fibrillar proteins, glycosaminoglycans, proteoglycans, minerals, and related proteins.^58^ *HSPG2*, *FBN1*, and *FBN2*, which were identified as candidate ECM genes in our patient cohort, have also been reported to be associated with AIS in specific cohorts,^43,44^ and several collagen genes (*COL6A3*, *COL11A2*, *COL5A1*, *COL11A1*) were also identified in this cohort. Previous researchers have suggested that the main pathogenesis of AIS is asymmetrical bone growth,^59^ and histological studies have shown decreased chondrogenesis, disorganized columnation, and premature cessation of growth in the cartilaginous growth plate of the vertebral body.^60^ The researchers proposed that growth plate chondrocytes indirectly regulate cellular activities, such as shape and volume preferential changes, through alterations in ECM synthesis and degradation.^61^ These previous studies in individuals with AIS, coupled with our genetic findings, suggest that ECM organization genes may play an important role in the development of the abnormal spinal curvature.

Genes related to muscle contraction were another enriched category in our dataset. *MYH2*, *DES*, *RYR1*, *ACTN2*, *MYH7*, and *TPM3*, which are involved in muscle contraction, were identified in our AIS cohort. There have been various studies on the asymmetry of the paraspinalis muscles in patients with AIS and recent literature has reported dominance of electromyography activity on the convex side of the scoliotic curve.^62–64^ The genetic findings from our group and others are supportive of a functional role for muscle contraction in AIS etiology. Although the pathogenesis of AIS is still not fully understood, we speculate that the asymmetry of the activity of structures involved in paraspinal muscle contraction as well as the ECM comprising the growth plate of the vertebral body are intricately connected within musculoskeletal tissues and are involved in the biomechanical system, which is one of the important etiologies of AIS.

The majority of previous research on the genetic basis of AIS were genome-wide association studies, which identified a number of SNPs associated with AIS. Although we had a small cohort, we were able to leverage the ethnic background of our participants to replicate the association of 4 known AIS SNPs. The rs12946942 locus at 17q24.3 is correlated with curve severity.^65^ It is located between *SOX9*, a pivotal transcription factor involved in skeletal development and ECM remodeling,^66,67^ and *KCNJ2*, which encodes a potassium channel involved in muscle contraction, and associated with skeletal malformations including progressive scoliosis.^68,69^ The rs7633294 at 3p14.1 is located near *MAGI1*, which encodes a scaffolding molecule important for the stabilization of cadherin-mediated cell interactions.^69,70,70^ The loci rs10756785 and rs3904778 are located within *BNC2*, which encodes a highly conserved zinc-finger protein involved in ECM regulation.^37,71–73^ Additionally, we detected 3 rare nonsynonymous variants with predicted benign effect in affected individuals, while no rare nonsynonymous variants were detected in unaffected individuals. These findings further support that the regulation of ECM structure and muscle contraction through transcriptional mechanisms play a significant role in AIS. Given the delicacy of these systems, identifying functional regulatory variants will be important to understand the precise mechanisms that lead to disordered phenotypes. Continued efforts in utilizing WGS combined with regulatory variant annotation have been successful in identifying candidate causative variants in other complex disorders like cardiomyopathy and autism spectrum disorder,^74,75^ and similar approaches should be employed to better understand the etiology of AIS.

In our study, 6% of all variants were identified as intronic variants and 4% as SVs as the causative variants of AIS. The identification of intronic variants and SVs require sequencing of noncoding regions of the genome. The panel-based sequencing and WES are limited in their ability to detect such variants, particularly those with breakpoints in intronic or intergenic regions, whereas WGS can overcome this limitation by covering all exonic and intronic regions of the genome.^76,77^ We believe that genetic approaches using WGS in these population will help us to understand the different phenotypes of patients in clinical practice and to characterize the genetic components of AIS pathophysiology. It is noteworthy that a variety of syndromic genes were identified in our patient cohort, suggesting that AIS may present as a milder presentation of more severe forms of syndromes. While many studies have reported on the important role of genetics in the initiation of AIS, it is also important to note that environmental factors and epigenetic influences are also deeply involved in the progression of AIS.^7,78^

This study had some limitations. The study population was predominantly of European and American ancestry. To capture the complete genetic landscape of AIS, WGS studies of larger cohorts including individuals with diverse ancestry will be required. Secondly, the majority of our identified variants are heterozygous, and without functional testing, it is unclear if these variants have a true dominant-negative effect on the resulting protein. Additional functional studies of specific variants will be warranted to demonstrate causality. Thirdly, analyses of clinical phenotypes of AIS, such as Cobb angle and location of the curve, are lacking. Further studies to determine the association between clinical manifestations and genotype will provide a deeper understanding of the role of genetic mechanisms underlying AIS. However, despite these limitations, the present study has the strength of being the first to investigate WGS results of the largest AIS cohort.

In conclusion, this study investigated the genetic landscape in the largest AIS cohort utilizing WGS. We observed an overrepresentation of variants in ECM organization and muscle contraction categories, with few specific genes shared across families. Overall, this study revealed that the genetic etiology of AIS is highly polygenic and that WGS has a greater potential to detect a wider spectrum of genetic variants, such as SVs and intronic variants. Further research with clinical phenotypes and functional studies of genetic variants are required to draw definitive conclusions regarding the genetic mechanisms in AIS pathogenesis.

## Data and code availability

The data presented in this study are available on request from the corresponding authors. The data are not publicly available due to the ethical and privacy nature of the data. The RareVision™ algorithm and associated in-house scripts are proprietary to Inocras Inc.

## Data Availability

The data presented in this study are available on request from the corresponding authors. The data are not publicly available due to the ethical and privacy nature of the data.

## Acknowledgments

This study was supported by a research grant of the Basic Science Research Program through the National Research Foundation of Korea (NRF) funded by the Ministry of Science and ICT (Information and Communication Technology, NRF-2021R1F1A1045417). The sponsor or funding organization had no role in the design or conduct of this research.

## Author contributions

Concept and design: I.O.T., E.K.L., S.L., K.S.; acquisition, analysis, and interpretation of data: I.O.T., E.K.L., A.G.; drafting of the manuscript: I.O.T., E.K.L.; critical revision of the manuscript for important intellectual content: S.L., K.S.; statistical analysis: I.O.T., E.K.L.; funding acquisition: E.K.L., S.L.; administrative, technical, and material support: Y.L., J-Y.K., W.L., A.G.; supervision: S.L., K.S.

## REFERENCES

1. Rogala, E.J., Drummond, D.S., and Gurr, J. (1978). Scoliosis: incidence and natural history. A prospective epidemiological study. J Bone Joint Surg Am 60, 173–176.

2. Asher, M.A., and Burton, D.C. (2006). Adolescent idiopathic scoliosis: natural history and long term treatment effects. Scoliosis 1, 2. 10.1186/1748-7161-1-2.

3. Soucacos, P.N., Soucacos, P.K., Zacharis, K.C., Beris, A.E., and Xenakis, T.A. (1997). School-screening for scoliosis. A prospective epidemiological study in northwestern and central Greece. J Bone Joint Surg Am 79, 1498–1503. 10.2106/00004623-199710000-00006.

4. Weinstein, S.L., Dolan, L.A., Cheng, J.C., Danielsson, A., and Morcuende, J.A. (2008). Adolescent idiopathic scoliosis. Lancet 371, 1527–1537. 10.1016/s0140-6736(08)60658-3.

5. Dunn, J., Henrikson, N.B., Morrison, C.C., Blasi, P.R., Nguyen, M., and Lin, J.S. (2018). Screening for Adolescent Idiopathic Scoliosis: Evidence Report and Systematic Review for the US Preventive Services Task Force. Jama 319, 173–187. 10.1001/jama.2017.11669.

6. Barton, C.B., and Weinstein, S.L. (2018). Adolescent Idiopathic Scoliosis: Natural History. In Pathogenesis of Idiopathic Scoliosis, M. Machida, S.L. Weinstein, and J. Dubousset, eds. (Springer Japan), pp. 27–50. 10.1007/978-4-431-56541-3_2.

7. Pérez-Machado, G., Berenguer-Pascual, E., Bovea-Marco, M., Rubio-Belmar, P.A., García-López, E., Garzón, M.J., Mena-Mollá, S., Pallardó, F.V., Bas, T., Viña, J.R., and García-Giménez, J.L. (2020). From genetics to epigenetics to unravel the etiology of adolescent idiopathic scoliosis. Bone 140, 115563. 10.1016/j.bone.2020.115563.

8. Tang, N.L., Yeung, H.Y., Hung, V.W., Di Liao, C., Lam, T.P., Yeung, H.M., Lee, K.M., Ng, B.K., and Cheng, J.C. (2012). Genetic epidemiology and heritability of AIS: A study of 415 Chinese female patients. J Orthop Res 30, 1464–1469. 10.1002/jor.22090.

9. Watanabe, K., Michikawa, T., Yonezawa, I., Takaso, M., Minami, S., Soshi, S., Tsuji, T., Okada, E., Abe, K., Takahashi, M., et al. (2017). Physical Activities and Lifestyle Factors Related to Adolescent Idiopathic Scoliosis. JBJS 99, 284–294. 10.2106/jbjs.16.00459.

10. Terhune, E.A., Wethey, C.I., Cuevas, M.T., Monley, A.M., Baschal, E.E., Bland, M.R., Baschal, R., Trahan, G.D., Taylor, M.R.G., Jones, K.L., and Hadley Miller, N. (2021). Whole Exome Sequencing of 23 Multigeneration Idiopathic Scoliosis Families Reveals Enrichments in Cytoskeletal Variants, Suggests Highly Polygenic Disease. Genes (Basel) 12. 10.3390/genes12060922.

11. Xia, C., Xu, L., Xue, B., Sheng, F., Qiu, Y., and Zhu, Z. (2019). Rare variant of HSPG2 is not involved in the development of adolescent idiopathic scoliosis: evidence from a large-scale replication study. BMC Musculoskelet Disord 20, 24. 10.1186/s12891-019-2402-x.

12. Takahashi, Y., Matsumoto, M., Karasugi, T., Watanabe, K., Chiba, K., Kawakami, N., Tsuji, T., Uno, K., Suzuki, T., Ito, M., et al. (2011). Replication study of the association between adolescent idiopathic scoliosis and two estrogen receptor genes. J Orthop Res 29, 834–837. 10.1002/jor.21322.

13. Sharma, S., Gao, X., Londono, D., Devroy, S.E., Mauldin, K.N., Frankel, J.T., Brandon, J.M., Zhang, D., Li, Q.-Z., Dobbs, M.B., et al. (2011). Genome-wide association studies of adolescent idiopathic scoliosis suggest candidate susceptibility genes. Human Molecular Genetics 20, 1456–1466. 10.1093/hmg/ddq571.

14. Sharma, S., Londono, D., Eckalbar, W.L., Gao, X., Zhang, D., Mauldin, K., Kou, I., Takahashi, A., Matsumoto, M., Kamiya, N., et al. (2015). A PAX1 enhancer locus is associated with susceptibility to idiopathic scoliosis in females. Nature Communications 6, 6452. 10.1038/ncomms7452.

15. Zhu, Z., Tang, N.L.-S., Xu, L., Qin, X., Mao, S., Song, Y., Liu, L., Li, F., Liu, P., Yi, L., et al. (2015). Genome-wide association study identifies new susceptibility loci for adolescent idiopathic scoliosis in Chinese girls. Nature Communications 6, 8355. 10.1038/ncomms9355.

16. Zhu, Z., Xu, L., Leung-Sang Tang, N., Qin, X., Feng, Z., Sun, W., Zhu, W., Shi, B., Liu, P., Mao, S., et al. (2017). Genome-wide association study identifies novel susceptible loci and highlights Wnt/beta-catenin pathway in the development of adolescent idiopathic scoliosis. Human Molecular Genetics 26, 1577–1583. 10.1093/hmg/ddx045.

17. Takahashi, Y., Kou, I., Takahashi, A., Johnson, T.A., Kono, K., Kawakami, N., Uno, K., Ito, M., Minami, S., Yanagida, H., et al. (2011). A genome-wide association study identifies common variants near LBX1 associated with adolescent idiopathic scoliosis. Nature Genetics 43, 1237–1240. 10.1038/ng.974.

18. Liu, S., Wu, N., Zuo, Y., Zhou, Y., Liu, J., Liu, Z., Chen, W., Liu, G., Chen, Y., Chen, J., et al. (2017). Genetic Polymorphism of LBX1 Is Associated With Adolescent Idiopathic Scoliosis in Northern Chinese Han Population. Spine 42, 1125–1129. 10.1097/brs.0000000000002111.

19. Liu, G., Liu, S., Lin, M., Li, X., Chen, W., Zuo, Y., Liu, J., Niu, Y., Zhao, S., Long, B., et al. (2018). Genetic polymorphisms of GPR126 are functionally associated with PUMC classifications of adolescent idiopathic scoliosis in a Northern Han population. Journal of Cellular and Molecular Medicine 22, 1964–1971. 10.1111/jcmm.13486.

20. Liu, G., Liu, S., Li, X., Chen, J., Chen, W., Zuo, Y., Liu, J., Niu, Y., Lin, M., Zhao, S., et al. (2019). Genetic polymorphisms of PAX1 are functionally associated with different PUMC types of adolescent idiopathic scoliosis in a northern Chinese Han population. Gene 688, 215–220. 10.1016/j.gene.2018.12.013.

21. Man, G.C.-W., Tang, N.L.-S., Chan, T.F., Lam, T.P., Li, J.W., Ng, B.K.-W., Zhu, Z., Qiu, Y., and Cheng, J.C.-Y. (2019). Replication Study for the Association of GWAS-associated Loci With Adolescent Idiopathic Scoliosis Susceptibility and Curve Progression in a Chinese Population. Spine 44, 464–471. 10.1097/brs.0000000000002866.

22. Qin, X., Xu, L., Xia, C., Zhu, W., Sun, W., Liu, Z., Qiu, Y., and Zhu, Z. (2017). Genetic Variant of GPR126 Gene is Functionally Associated With Adolescent Idiopathic Scoliosis in Chinese Population. Spine 42, E1098–E1103. 10.1097/brs.0000000000002123.

23. Xu, L., Sheng, F., Xia, C., Qin, X., Tang, N.L.-S., Qiu, Y., Cheng, J.C.-Y., and Zhu, Z. (2018). Genetic Variant of PAX1 Gene Is Functionally Associated With Adolescent Idiopathic Scoliosis in the Chinese Population. Spine 43, 492–496. 10.1097/brs.0000000000002475.

24. Xu, L., Xia, C., Qin, X., Sun, W., Tang, N.L.-S., Qiu, Y., Cheng, J.C.-Y., and Zhu, Z. (2017). Genetic variant of BNC2 gene is functionally associated with adolescent idiopathic scoliosis in Chinese population. Molecular Genetics and Genomics 292, 789–794. 10.1007/s00438-017-1315-3.

25. Wu, Z., Dai, Z., Yuwen, W., Liu, Z., Qiu, Y., Cheng, J.C.-Y., Zhu, Z., and Xu, L. (2021). Genetic Variants of CHD7 Are Associated with Adolescent Idiopathic Scoliosis. Spine 46, E618–E624. 10.1097/brs.0000000000003857.

26. Li, Y., Wu, Z., Xu, L., Feng, Z., Wang, Y., Dai, Z., Liu, Z., Sun, X., Qiu, Y., and Zhu, Z. (2021). Genetic Variant of TBX1 Gene Is Functionally Associated With Adolescent Idiopathic Scoliosis in the Chinese Population. Spine 46, 17–21. 10.1097/brs.0000000000003700.

27. Grauers, A., Wang, J., Einarsdottir, E., Simony, A., Danielsson, A., Åkesson, K., Ohlin, A., Halldin, K., Grabowski, P., Tenne, M., et al. (2015). Candidate gene analysis and exome sequencing confirm <em>LBX1</em> as a susceptibility gene for idiopathic scoliosis. The Spine Journal 15, 2239–2246. 10.1016/j.spinee.2015.05.013.

28. Nada, D., Julien, C., Samuels, M.E., and Moreau, A. (2018). A Replication Study for Association of LBX1 Locus With Adolescent Idiopathic Scoliosis in French–Canadian Population. Spine 43, 172–178. 10.1097/brs.0000000000002280.

29. Pennisi, E. (2022). Upstart DNA sequencers could be a ‘game changer’. Science 376, 1257–1258. 10.1126/science.add4867.

30. Horne, J.P., Flannery, R., and Usman, S. (2014). Adolescent idiopathic scoliosis: diagnosis and management. Am Fam Physician 89, 193–198.

31. Auton, A., Brooks, L.D., Durbin, R.M., Garrison, E.P., Kang, H.M., Korbel, J.O., Marchini, J.L., McCarthy, S., McVean, G.A., and Abecasis, G.R. (2015). A global reference for human genetic variation. Nature 526, 68–74. 10.1038/nature15393.

32. Karczewski, K.J., Francioli, L.C., Tiao, G., Cummings, B.B., Alföldi, J., Wang, Q., Collins, R.L., Laricchia, K.M., Ganna, A., Birnbaum, D.P., et al. (2020). The mutational constraint spectrum quantified from variation in 141,456 humans. Nature 581, 434–443. 10.1038/s41586-020-2308-7.

33. Karczewski, K.J., Weisburd, B., Thomas, B., Solomonson, M., Ruderfer, D.M., Kavanagh, D., Hamamsy, T., Lek, M., Samocha, K.E., Cummings, B.B., et al. (2017). The ExAC browser: displaying reference data information from over 60 000 exomes. Nucleic Acids Res 45, D840–d845. 10.1093/nar/gkw971.

34. Richards, S., Aziz, N., Bale, S., Bick, D., Das, S., Gastier-Foster, J., Grody, W.W., Hegde, M., Lyon, E., Spector, E., et al. (2015). Standards and guidelines for the interpretation of sequence variants: a joint consensus recommendation of the American College of Medical Genetics and Genomics and the Association for Molecular Pathology. Genet Med 17, 405–424. 10.1038/gim.2015.30.

35. Kou, I., Takahashi, Y., Johnson, T.A., Takahashi, A., Guo, L., Dai, J., Qiu, X., Sharma, S., Takimoto, A., Ogura, Y., et al. (2013). Genetic variants in GPR126 are associated with adolescent idiopathic scoliosis. Nat Genet 45, 676–679. 10.1038/ng.2639.

36. Li, W., Li, Y., Zhang, L., Guo, H., Tian, D., Li, Y., Peng, Y., Zheng, Y., Dai, Y., Xia, K., et al. (2016). AKAP2 identified as a novel gene mutated in a Chinese family with adolescent idiopathic scoliosis. J Med Genet 53, 488–493. 10.1136/jmedgenet-2015-103684.

37. Ogura, Y., Kou, I., Miura, S., Takahashi, A., Xu, L., Takeda, K., Takahashi, Y., Kono, K., Kawakami, N., Uno, K., et al. (2015). A Functional SNP in BNC2 Is Associated with Adolescent Idiopathic Scoliosis. Am J Hum Genet 97, 337–342. 10.1016/j.ajhg.2015.06.012.

38. Gao, X., Gordon, D., Zhang, D., Browne, R., Helms, C., Gillum, J., Weber, S., Devroy, S., Swaney, S., Dobbs, M., et al. (2007). CHD7 gene polymorphisms are associated with susceptibility to idiopathic scoliosis. Am J Hum Genet 80, 957–965. 10.1086/513571.

39. Tilley, M.K., Justice, C.M., Swindle, K., Marosy, B., Wilson, A.F., and Miller, N.H. (2013). CHD7 gene polymorphisms and familial idiopathic scoliosis. Spine (Phila Pa 1976) 38, E1432–1436. 10.1097/BRS.0b013e3182a51781.

40. Yu, H., Khanshour, A.M., Ushiki, A., Otomo, N., Koike, Y., Einarsdottir, E., Fan, Y., Antunes, L., Kidane, Y.H., Cornelia, R., et al. (2024). Association of genetic variation in COL11A1 with adolescent idiopathic scoliosis. Elife 12. 10.7554/eLife.89762.

41. Rebello, D., Wohler, E., Erfani, V., Li, G., Aguilera, A.N., Santiago-Cornier, A., Zhao, S., Hwang, S.W., Steiner, R.D., Zhang, T.J., et al. (2023). COL11A2 as a candidate gene for vertebral malformations and congenital scoliosis. Hum Mol Genet 32, 2913–2928. 10.1093/hmg/ddad117.

42. Nada, D., Julien, C., Papillon-Cavanagh, S., Majewski, J., Elbakry, M., Elremaly, W., Samuels, M.E., and Moreau, A. (2022). Identification of FAT3 as a new candidate gene for adolescent idiopathic scoliosis. Sci Rep 12, 12298. 10.1038/s41598-022-16620-6.

43. Buchan, J.G., Alvarado, D.M., Haller, G.E., Cruchaga, C., Harms, M.B., Zhang, T., Willing, M.C., Grange, D.K., Braverman, A.C., Miller, N.H., et al. (2014). Rare variants in FBN1 and FBN2 are associated with severe adolescent idiopathic scoliosis. Hum Mol Genet 23, 5271–5282. 10.1093/hmg/ddu224.

44. Baschal, E.E., Wethey, C.I., Swindle, K., Baschal, R.M., Gowan, K., Tang, N.L., Alvarado, D.M., Haller, G.E., Dobbs, M.B., Taylor, M.R., et al. (2014). Exome sequencing identifies a rare HSPG2 variant associated with familial idiopathic scoliosis. G3 (Bethesda) 5, 167–174. 10.1534/g3.114.015669.

45. Terhune, E.A., Cuevas, M.T., Monley, A.M., Wethey, C.I., Chen, X., Cattell, M.V., Bayrak, M.N., Bland, M.R., Sutphin, B., Trahan, G.D., et al. (2021). Mutations in KIF7 implicated in idiopathic scoliosis in humans and axial curvatures in zebrafish. Hum Mutat 42, 392–407. 10.1002/humu.24162.

46. Jennings, W., Hou, M., Perterson, D., Missiuna, P., Thabane, L., Tarnopolsky, M., and Samaan, M.C. (2019). Paraspinal muscle ladybird homeobox 1 (LBX1) in adolescent idiopathic scoliosis: a cross-sectional study. Spine J 19, 1911–1916. 10.1016/j.spinee.2019.06.014.

47. Hassan, A., Parent, S., Mathieu, H., Zaouter, C., Molidperee, S., Bagu, E.T., Barchi, S., Villemure, I., Patten, S.A., and Moldovan, F. (2019). Adolescent idiopathic scoliosis associated POC5 mutation impairs cell cycle, cilia length and centrosome protein interactions. PLoS One 14, e0213269. 10.1371/journal.pone.0213269.

48. Purcell, S., Neale, B., Todd-Brown, K., Thomas, L., Ferreira, M.A., Bender, D., Maller, J., Sklar, P., de Bakker, P.I., Daly, M.J., and Sham, P.C. (2007). PLINK: a tool set for whole-genome association and population-based linkage analyses. Am J Hum Genet 81, 559–575. 10.1086/519795.

49. Chang, C.C., Chow, C.C., Tellier, L.C., Vattikuti, S., Purcell, S.M., and Lee, J.J. (2015). Second-generation PLINK: rising to the challenge of larger and richer datasets. Gigascience 4, 7. 10.1186/s13742-015-0047-8.

50. Kanehisa, M., and Goto, S. (2000). KEGG: kyoto encyclopedia of genes and genomes. Nucleic Acids Res 28, 27–30. 10.1093/nar/28.1.27.

51. Ashburner, M., Ball, C.A., Blake, J.A., Botstein, D., Butler, H., Cherry, J.M., Davis, A.P., Dolinski, K., Dwight, S.S., Eppig, J.T., et al. (2000). Gene Ontology: tool for the unification of biology. Nature Genetics 25, 25–29. 10.1038/75556.

52. Consortium, T.G.O. (2020). The Gene Ontology resource: enriching a GOld mine. Nucleic Acids Research 49, D325–D334. 10.1093/nar/gkaa1113.

53. Kuleshov, M.V., Jones, M.R., Rouillard, A.D., Fernandez, N.F., Duan, Q., Wang, Z., Koplev, S., Jenkins, S.L., Jagodnik, K.M., Lachmann, A., et al. (2016). Enrichr: a comprehensive gene set enrichment analysis web server 2016 update. Nucleic Acids Res 44, W90–97. 10.1093/nar/gkw377.

54. Szklarczyk, D., Kirsch, R., Koutrouli, M., Nastou, K., Mehryary, F., Hachilif, R., Gable, A.L., Fang, T., Doncheva, N.T., Pyysalo, S., et al. (2023). The STRING database in 2023: protein-protein association networks and functional enrichment analyses for any sequenced genome of interest. Nucleic Acids Res 51, D638–d646. 10.1093/nar/gkac1000.

55. Morava, E., Lacassie, Y., King, A., Illes, T., and Marble, M. (2002). Scoliosis in velo-cardio-facial syndrome. J Pediatr Orthop 22, 780–783.

56. Du, Q., de la Morena, M.T., and van Oers, N.S.C. (2019). The Genetics and Epigenetics of 22q11.2 Deletion Syndrome. Front Genet 10, 1365. 10.3389/fgene.2019.01365.

57. Wiel, L.C., Bruno, I., Barbi, E., and Sirchia, F. (2022). From Wolf-Hirschhorn syndrome to NSD2 haploinsufficiency: a shifting paradigm through the description of a new case and a review of the literature. Ital J Pediatr 48, 72. 10.1186/s13052-022-01267-w.

58. Lamandé, S.R., and Bateman, J.F. (2020). Genetic Disorders of the Extracellular Matrix. Anat Rec (Hoboken) 303, 1527–1542. 10.1002/ar.24086.

59. Michelsson, J.E. (1965). The development of spinal deformity in experimental scoliosis. Acta Orthop Scand Suppl, Suppl 81:81–91. 10.3109/ort.1965.36.suppl-81.01.

60. Stilwell, D.L., Jr. (1962). Structural deformities of vertebrae. Bone adaptation and modeling in experimental scoliosis and kyphosis. J Bone Joint Surg Am 44-a, 611–634.

61. Wang, S., Qiu, Y., Ma, Z., Xia, C., Zhu, F., and Zhu, Z. (2010). Expression of Runx2 and type X collagen in vertebral growth plate of patients with adolescent idiopathic scoliosis. Connect Tissue Res 51, 188–196. 10.3109/03008200903215590.

62. Cheung, J., Veldhuizen, A.G., Halbertsma, J.P., Maurits, N.M., Sluiter, W.J., Cool, J.C., and Van Horn, J.R. (2004). The relation between electromyography and growth velocity of the spine in the evaluation of curve progression in idiopathic scoliosis. Spine (Phila Pa 1976) 29, 1011–1016. 10.1097/00007632-200405010-00012.

63. Farahpour, N., Ghasemi, S., Allard, P., and Saba, M.S. (2014). Electromyographic responses of erector spinae and lower limb’s muscles to dynamic postural perturbations in patients with adolescent idiopathic scoliosis. J Electromyogr Kinesiol 24, 645–651. 10.1016/j.jelekin.2014.05.014.

64. Stetkarova, I., Zamecnik, J., Bocek, V., Vasko, P., Brabec, K., and Krbec, M. (2016). Electrophysiological and histological changes of paraspinal muscles in adolescent idiopathic scoliosis. Eur Spine J 25, 3146–3153. 10.1007/s00586-016-4628-8.

65. Miyake, A., Kou, I., Takahashi, Y., Johnson, T.A., Ogura, Y., Dai, J., Qiu, X., Takahashi, A., Jiang, H., Yan, H., et al. (2013). Identification of a susceptibility locus for severe adolescent idiopathic scoliosis on chromosome 17q24.3. PLoS One 8, e72802. 10.1371/journal.pone.0072802.

66. Tsingas, M., Ottone, O.K., Haseeb, A., Barve, R.A., Shapiro, I.M., Lefebvre, V., and Risbud, M.V. (2020). Sox9 deletion causes severe intervertebral disc degeneration characterized by apoptosis, matrix remodeling, and compartment-specific transcriptomic changes. Matrix Biol 94, 110–133. 10.1016/j.matbio.2020.09.003.

67. Güven, A., Kalebic, N., Long, K.R., Florio, M., Vaid, S., Brandl, H., Stenzel, D., and Huttner, W.B. (2020). Extracellular matrix-inducing Sox9 promotes both basal progenitor proliferation and gliogenesis in developing neocortex. Elife 9. 10.7554/eLife.49808.

68. MJCMD, V. (2015). Genes associated with adolescent idiopathic scoliosis: a review. Hereditary Genet 4, 2161–1041.1000146.

69. Blyth, M., Huang, S., Maloney, V., Crolla, J.A., and Karen Temple, I. (2008). A 2.3Mb deletion of 17q24.2-q24.3 associated with ‘Carney Complex plus’. Eur J Med Genet 51, 672–678. 10.1016/j.ejmg.2008.09.002.

70. Mizuhara, E., Nakatani, T., Minaki, Y., Sakamoto, Y., Ono, Y., and Takai, Y. (2005). MAGI1 recruits Dll1 to cadherin-based adherens junctions and stabilizes it on the cell surface. J Biol Chem 280, 26499–26507. 10.1074/jbc.M500375200.

71. Khanshour, A.M., Kou, I., Fan, Y., Einarsdottir, E., Makki, N., Kidane, Y.H., Kere, J., Grauers, A., Johnson, T.A., Paria, N., et al. (2018). Genome-wide meta-analysis and replication studies in multiple ethnicities identify novel adolescent idiopathic scoliosis susceptibility loci. Hum Mol Genet 27, 3986–3998. 10.1093/hmg/ddy306.

72. Bobowski-Gerard, M., Boulet, C., Zummo, F.P., Dubois-Chevalier, J., Gheeraert, C., Bou Saleh, M., Strub, J.M., Farce, A., Ploton, M., Guille, L., et al. (2022). Functional genomics uncovers the transcription factor BNC2 as required for myofibroblastic activation in fibrosis. Nat Commun 13, 5324. 10.1038/s41467-022-33063-9.

73. Orang, A., Dredge, B.K., Liu, C.Y., Bracken, J.M., Chen, C.H., Sourdin, L., Whitfield, H.J., Lumb, R., Boyle, S.T., Davis, M.J., et al. (2023). Basonuclin-2 regulates extracellular matrix production and degradation. Life Sci Alliance 6. 10.26508/lsa.202301984.

74. Tuncay, I.O., Parmalee, N.L., Khalil, R., Kaur, K., Kumar, A., Jimale, M., Howe, J.L., Goodspeed, K., Evans, P., Alzghoul, L., et al. (2022). Analysis of recent shared ancestry in a familial cohort identifies coding and noncoding autism spectrum disorder variants. NPJ Genom Med 7, 13. 10.1038/s41525-022-00284-2.

75. Lesurf, R., Said, A., Akinrinade, O., Breckpot, J., Delfosse, K., Liu, T., Yao, R., Persad, G., McKenna, F., Noche, R.R., et al. (2022). Whole genome sequencing delineates regulatory, copy number, and cryptic splice variants in early onset cardiomyopathy. NPJ Genom Med 7, 18. 10.1038/s41525-022-00288-y.

76. Meynert, A.M., Ansari, M., FitzPatrick, D.R., and Taylor, M.S. (2014). Variant detection sensitivity and biases in whole genome and exome sequencing. BMC Bioinformatics 15, 247. 10.1186/1471-2105-15-247.

77. Belkadi, A., Bolze, A., Itan, Y., Cobat, A., Vincent, Q.B., Antipenko, A., Shang, L., Boisson, B., Casanova, J.L., and Abel, L. (2015). Whole-genome sequencing is more powerful than whole-exome sequencing for detecting exome variants. Proc Natl Acad Sci U S A 112, 5473–5478. 10.1073/pnas.1418631112.

78. Goldberg, C.J., Dowling, F.E., and Fogarty, E.E. (1993). Adolescent idiopathic scoliosis: is rising growth rate the triggering factor in progression? Eur Spine J 2, 29–36. 10.1007/bf00301052.

