## Supplementary data for "Whole Genome Sequencing in Adolescent Idiopathic Scoliosis Cohort Indicates Polygenic Disease Involving Multiple Biological Pathways"

**Supplementary Table**

**Supplementary Table 1. Number of rare nonsynonymous point mutations**

| **Gene** | **All** | | **Benign / Likely Benign** | | **PTV, Pathogenic / Likely Pathogenic, or VUS** | |
| --- | --- | --- | --- | --- | --- | --- |
|  | **Affected** | **Unaffected** | **Affected** | **Unaffected** | **Affected** | **Unaffected** |
| *BNC2* | 3 | 0 | 3 | 0 | 0 | 0 |
| *KIF7* | 4 | 0 | 3 | 0 | 1 | 0 |
| *COL11A1* | 4 | 1 | 2 | 0 | 2 | 1 |
| *COL11A2* | 6 | 3 | 4 | 2 | 2 | 1 |
| *FBN1* | 4 | 2 | 2 | 0 | 2 | 2 |
| *HSPG2* | 7 | 5 | 4 | 3 | 3 | 2 |
| *POC5* | 2 | 1 | 1 | 0 | 1 | 1 |
| *FBN2* | 3 | 3 | 0 | 1 | 3 | 2 |
| *ADGRG6* | 0 | 1 | 0 | 1 | 0 | 0 |
| *FAT3* | 2 | 9 | 1 | 4 | 1 | 5 |

**Supplementary Figure**

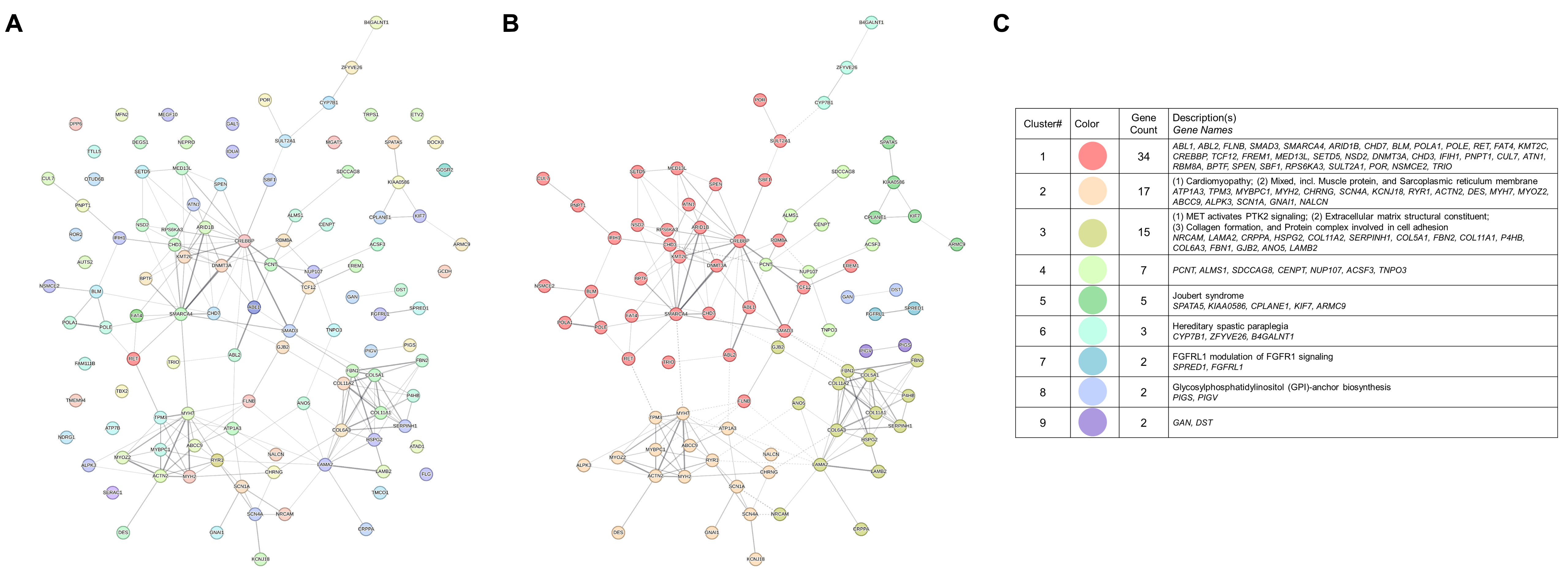

**Supplementary Figure 1.** **STRING functional interaction map of the candidate genes**

Analysis was done using default settings. Line thickness indicates the strength of data support. **(A)** Non-clustered interaction map of all 137 candidate genes. **(B)** Clustered interaction map. Disconnected nodes were excluded, leaving 87 genes. K-means clustering was done with K=5 through K=12, then K=9 was selected as the number of clusters using the elbow (data now shown). **(C)** Legend for the clusters shown in (B).
